# Plasma p-tau217 and NfL predict incident dementia in the community

**DOI:** 10.64898/2026.08.07.26359934

**Authors:** Anita Lenora Sunde, Diego A Tovar-Rios, Audun Osland Vik-Mo, Henrik Zetterberg, Burak Arslan, Kübra Tan, Hanna Huber, Karin Persson, Guglielmo Di Molfetta, Ilaria Pola, Marit Næss, Håvard K Skjellegrind, Geir Selbaek, Nicholas J Ashton, Dag Aarsland

**Affiliations:** Centre for Age-Related Medicine (SESAM), Stavanger University Hospital, Stavanger, Norway; Department of Clinical Medicine, University of Bergen, Bergen, Norway; L-BioStat, KU Leuven, Leuven, Belgium; Grupo de Investigación en Estadística Aplicada-INFERIR, Universidad del Valle, Santiago de Cali, Colombia; Prevención y Control de la Enfermedad Crónica-PRECEC, Universidad del Valle, Santiago de Cali, Colombia; Department of Psychiatry and Neurochemistry, Institute of Neuroscience and Physiology, The Sahlgrenska Academy, University of Gothenburg, Gothenburg, Sweden; Department of Neurodegenerative Disease, UCL Institute of Neurology, London, UK; UK Dementia Research Institute at UCL, London, UK; Department of Pathology and Laboratory Medicine, University of Wisconsin School of Medicine and Public Health, Madison, Wisconsin, USA; Wisconsin Alzheimer’s Disease Research Center, University of Wisconsin School of Medicine and Public Health, Madison, Wisconsin, USA; Clinical Neurochemistry Laboratory, Sahlgrenska University Hospital, Gothenburg, Sweden; Paris Brain Institute, ICM, Pitié-Salpêtrière Hospital, Sorbonne University, Paris, France; Neurodegenerative Disorder Research Center, Division of Life Sciences and Medicine, and Department of Neurology, Institute on Aging and Brain Disorders, University of Science and Technology of China and First Affiliated Hospital of USTC, Hefei, P.R. China; German Center of Neurodegenerative Diseases (DZNE), Bonn, Germany; Department of Old Age Psychiatry and Cognitive Disorders, University Hospital Bonn, Bonn, Germany; Norwegian National Centre for Aging and Health, Vestfold Hospital Trust, Tønsberg, Norway; Department of Geriatric Medicine, Oslo University Hospital, Nydalen, Oslo, Norway; HUNT Research Centre, Department of Public Health and Nursing, NTNU, Norwegian University of Science and Technology, Levanger, Norway; Levanger Hospital, Nord-Trøndelag Hospital Trust, Levanger, Norway; Institute of Clinical Medicine, University of Oslo, Oslo, Norway; Banner Sun Health Research Institute, Sun City, Arizona, USA; Banner Alzheimer’s Institute and University of Arizona, Phoenix, Arizona, USA; Centre for Healthy Brain Ageing, Department of Psychological Medicine, Institute of Psychiatry, Psychology, and Neuroscience, King’s College London, London, UK

**Keywords:** Alzheimer’s disease pathology, dementia incidence, community-based, blood-based biomarkers, p-tau217, NfL, predictive performance, HUNT Study

## Abstract

**INTRODUCTION:** Characterizing the prognostic utility of blood-based biomarkers for Alzheimer’s disease (AD) in predicting longitudinal cognitive trajectories is essential; however, population-based evidence is needed.

**METHODS:** We evaluated plasma phosphorylated tau at threonine 217 (p-tau217) and plasma neurofilament light chain (NfL) in 4,971 dementia-free individuals aged 70 years and older from the population-based Norwegian HUNT study. Predefined cut-offs categorized biomarker ranges (p-tau217: low, intermediate, high; NfL: low, high), in addition to continuous biomarker analysis.

**RESULTS:** Adjusted for other risk factors, higher baseline p-tau217 and NfL ranges indicated a significantly increased dementia risk after four years compared to low ranges (intermediate p-tau217: risk ratio [RR] 1.23, 95% CI 1.01–1.50; high p-tau217: RR 2.05, 95% CI 1.73–2.44; high NfL: RR 1.72, 95% CI 1.31–2.27; jointly high p-tau217 and NfL: RR 3.32, 95% CI 2.61–4.23). The estimated cumulative risk of all-cause dementia was 10.6% (95% CI 9.3–12.1) for low p-tau217, 16.9% (95% CI 14.6-19.5) for intermediate p-tau217, 33.0% (95% CI 29.9–36.1) for high p-tau217, 15.7% (95% CI 14.4–17.0) for low NfL, 35.4% (95% CI 30.6–40.5) for high NfL, and 47.8% (95% CI 40.7–55.0) for jointly high p-tau217 and NfL. The association of p-tau217 with incident dementia differed by sex.

**DISCUSSION:** These findings support the use of blood-based biomarkers for population-level dementia risk stratification, underscore the value of combining markers to improve prognostic precision, and can aid clinicians using plasma p-tau217 or NfL in interpreting dementia risk.

## BACKGROUND

Alzheimer’s disease (AD) is the most common neurodegenerative disease, impacting millions of people worldwide.[1] The neuropathological changes of AD (ADNC) develop decades before clinical symptoms emerge, which are initially subtle before progressing via a prodromal stage of mild cognitive impairment (MCI) to dementia.[2–4] Knowing the risk factors for dementia is crucial for individual and societal planning. In addition to age, genetic, socioeconomic and lifestyle risk factors,[5] biomarkers might add to dementia prediction and treatment stratification. Blood concentrations of phosphorylated tau at threonine 217 (p-tau217) and neurofilament light chain (NfL) enable scalable assessments of ADNC and neurodegeneration,[6, 7] with the potential to identify AD pathology[8] and increased risk of cognitive decline among cognitively unimpaired (CU) or mild cognitively impaired older adults.[9–11] While plasma p-tau217 is specific for AD,[6] NfL is a general marker of neuro-axonal damage which can be seen in a range of neurological disorders.[12] Given limited evidence from population-based longitudinal studies,[13] we test the hypothesis that plasma p-tau217 and NfL are associated with 4-year incident dementia risk in individuals aged 70 years and older.

## METHODS

### Cohort

The Trøndelag Health Study (HUNT) is a population-based longitudinal health study conducted in Trøndelag county in Central Norway. The region’s key demographic and health indicators mirror Norwegian national averages closely. Starting in the 1980s, four study waves have been conducted to date, one every decade, and achieving high participation rates. The HUNT study uses a municipality-by-municipality data collection model, with local inclusion periods lasting a couple of weeks in the smallest municipalities to several months in the largest municipalities, before moving to the next municipality.[14] At the last study wave, HUNT4 (September 2017 to February 2019), all residents aged 70 years or older were invited to a standardized cognitive assessment (HUNT4 70+ substudy),[15] with a follow-up four years later (HUNT Aging in Trøndelag, AiT, September 2021 to June 2023).[16, 17] A detailed overview of included study participants is shown in Figure 1. This study was approved by the Regional Committee for Medical and Health Research Ethics in Norway (REC Southeast C 565876). Participation in HUNT required informed consent. In participants with reduced capacity to consent, next of kin gave consent.

**Figure 1.**
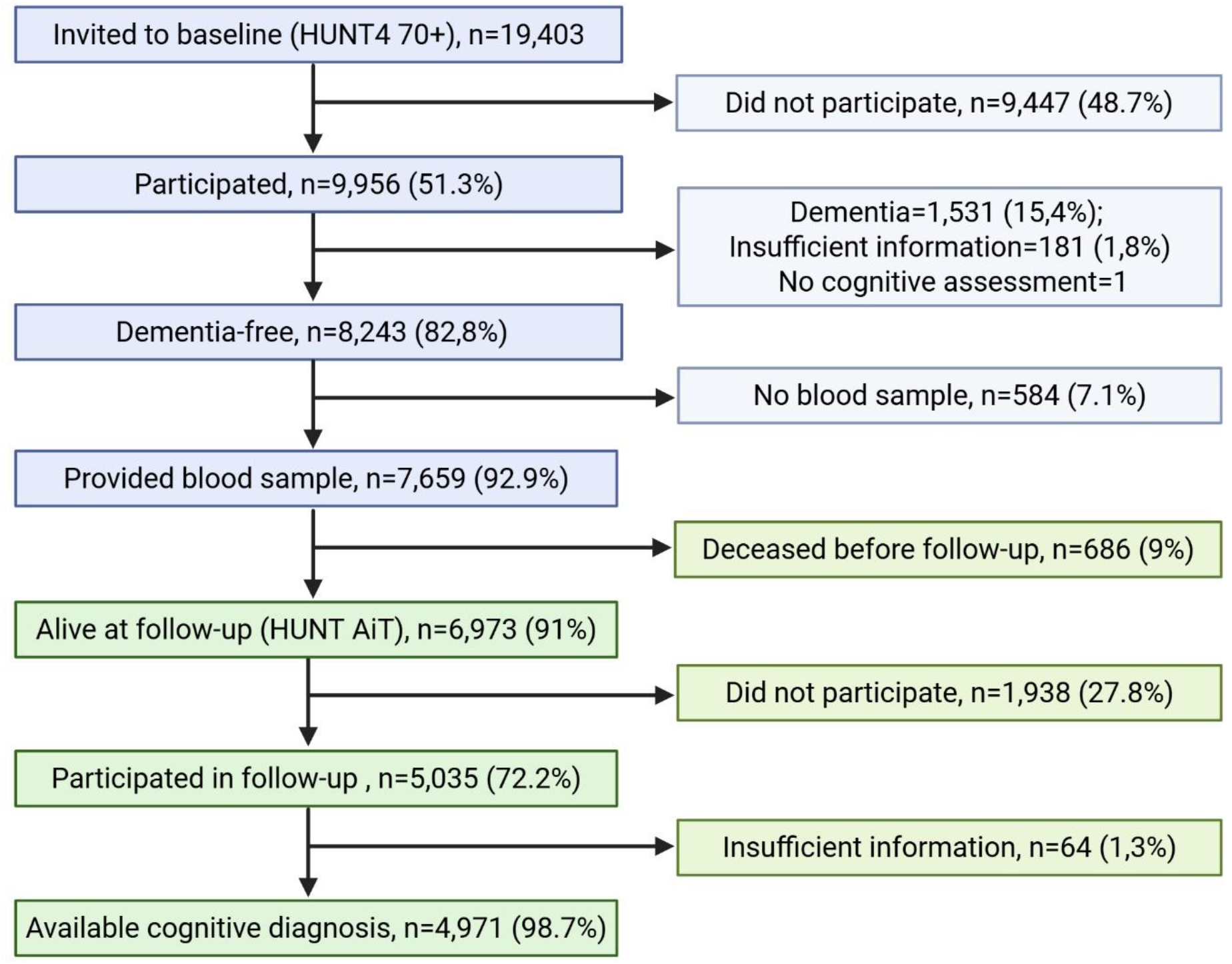
Overview of the study cohort. HUNT4 70+, The Trøndelag Health Study 4th wave 70+; AiT, The Trøndelag Health Study 4th wave Aging in Trøndelag.

### Cognitive assessment and diagnosis

At baseline and follow-up, health personnel assessed the participants’ cognitive, neuropsychiatric, and functional status using standardized interviews and clinical assessments at field stations, at homes, or in nursing homes, as described previously.[15–17] Physicians made diagnoses according to The Diagnostic and Statistical Manual of Mental Disorders, Fifth Edition (DSM-5) criteria (cognitively unimpaired (CU), MCI (minor neurocognitive disorder in DSM-5) or dementia (major neurocognitive disorder in DSM-5).[15–18] Biomarker data were not available when the clinical diagnoses were made, nor brain imaging data. Physicians assessing cognitive status at follow-up were blinded to baseline cognitive diagnoses. Details on the instruments used in the assessment can be found in the **Supplementary**

Apolipoprotein E (*APOE*) genotype and serum creatinine data were derived from the HUNT biobank.[17] Comorbidities and education levels were self-reported. Educational attainment was categorized into primary, secondary, and tertiary levels, defined as ≤10, 11-13, and ≥14 years of education, respectively. Body mass index (BMI, kg/m²) was categorized into underweight (<18.5 kg/m²), normal weight (18.5-24.9 kg/m²), overweight (25-29.9 kg/m²) and obesity (≥30 kg/m²).

### Plasma p-tau217 and NfL analysis

Blood samples were collected, handled, and stored according to standardized procedures.[19] Phlebotomy-to-freezing time and transportation ensured plasma p-tau217 and NfL stability.[20] Biomarkers were analyzed on the Simoa HD-X instrument (Quanterix, USA) at the University of Gothenburg, Sweden, with validated commercial kits (ALZpath p-Tau 217 Advantage PLUS, Quanterix; NfL Advantage V2, Quanterix). Using previously validated cut-offs, p-tau217 was categorized as low (<0.40 picograms per milliliter, pg/mL; 95% sensitivity for ADNC), intermediate (0.40–0.62 pg/mL), or high (≥0.63 pg/mL; 95% specificity for ADNC).[21, 22] Age-dependent NfL cut-offs were used,[23] with concentrations above the cut-off indicating neuroaxonal damage (70 years: 20pg/ml; ≥ 71 years: 35 pg/mL).[24] Details on blood sample handling procedures and a summary of the analytical performance of the assays can be found in the **Supplementary**.

### Statistical analysis

Group comparisons were performed using Kruskal–Wallis tests for continuous variables and Fisher’s exact tests for categorical variables. To account for differential participation and potential selection bias across study phases, inverse probability weighting (IPW) was incorporated in the inferential analysis. Weights calculation is described in detail in the **Supplementary**. Outcome was all-cause dementia at follow-up. Clinically diagnosed Alzheimer’s disease dementia (ADD) was described as a supplementary outcome (available in the **Supplementary**).

Distributions of plasma p-tau217 and NfL according to dementia outcomes were compared using Wilcoxon rank-sum tests and visualized using boxplots. Weighted cumulative absolute risk (AR) and risk ratios (RRs) for dementia were estimated using inverse probability-weighted proportions with corresponding 95% confidence intervals, stratified by baseline cognitive status (CU, MCI), sex, age group, *APOE* genotype, BMI and educational level. Associations between biomarker ranges and dementia outcomes were quantified using inverse probability-weighted generalized linear models with a quasi-Poisson distribution and log link to estimate RRs and 95% confidence intervals (CI). Models included interaction terms between biomarker categories and baseline cognitive status, and stratified analyses were conducted to evaluate effect modification by demographic and genetic factors. We explored the inclusion of time between baseline and follow-up in the models without finding a significant contribution. Pairwise contrasts between biomarker categories were estimated on the log scale and exponentiated to derive RRs. The presence of significant differences in RR between biomarker ranges across sex was evaluated by rate of risk ratios tests.

Odds ratios (OR) for all-cause dementia associated with continuous plasma p-tau217 and NfL concentrations, and clinical covariates (cognitive status, sex, age group, serum creatinine, *APOE* genotype, BMI and educational level) were explored with a survey-weighted logistic regression. Given the weighted cumulative incidence of dementia (17.7%), odds ratios may moderately overestimate risk ratios. A survey-weighted log-binomial model was explored but exhibited numerical instability; therefore, the logistic regression model was retained as the primary analysis. Continuous plasma p-tau217 was included linearly, and NfL with a second-order spline. Additional analyses assessed whether associations between plasma p-tau217 range, NfL range and dementia risk differed according to major baseline comorbidities (cerebrovascular disease, chronic obstructive pulmonary disease, cardiovascular disease, diabetes mellitus, and rheumatic disease). These models adjusted for age, sex, education, *APOE* genotype, serum creatinine and baseline cognitive status, and included interaction terms between biomarker ranges and each comorbidity.

All statistical analyses were performed using R 4.5.2. Statistical tests were two-sided, and a *P* value <.05 was considered statistically significant.

## RESULTS

In total, 4,971 participants (53.1% female, mean baseline age 75.8 ± 4.69 years, 36.2% baseline MCI) were dementia-free with an available blood sample at baseline and cognitive diagnosis at 4-year follow-up (mean follow-up time 4.21 ± SD 0.28 years; median 4.16, interquartile range 0.41 years). Of these, 695 (14%; IPW 17.7%, 95% CI 16.5-19.0) had developed dementia (Tables 1 and S1). Those who developed dementia were older, had less education, more often had the *APOE* ε4 allele and a lower baseline MoCA score than those without dementia at follow-up, while sex did not differ between groups. Dementia was more common among those with a diagnosis of MCI (25.6%) than those with CU (7.4%) at baseline. High range p-tau217 (n=1101) was more common than high range NfL (n=457). Median plasma p-tau217 and NfL concentration were higher in those who developed incident dementia compared to those who did not (Figure 2).

**Figure 2.**
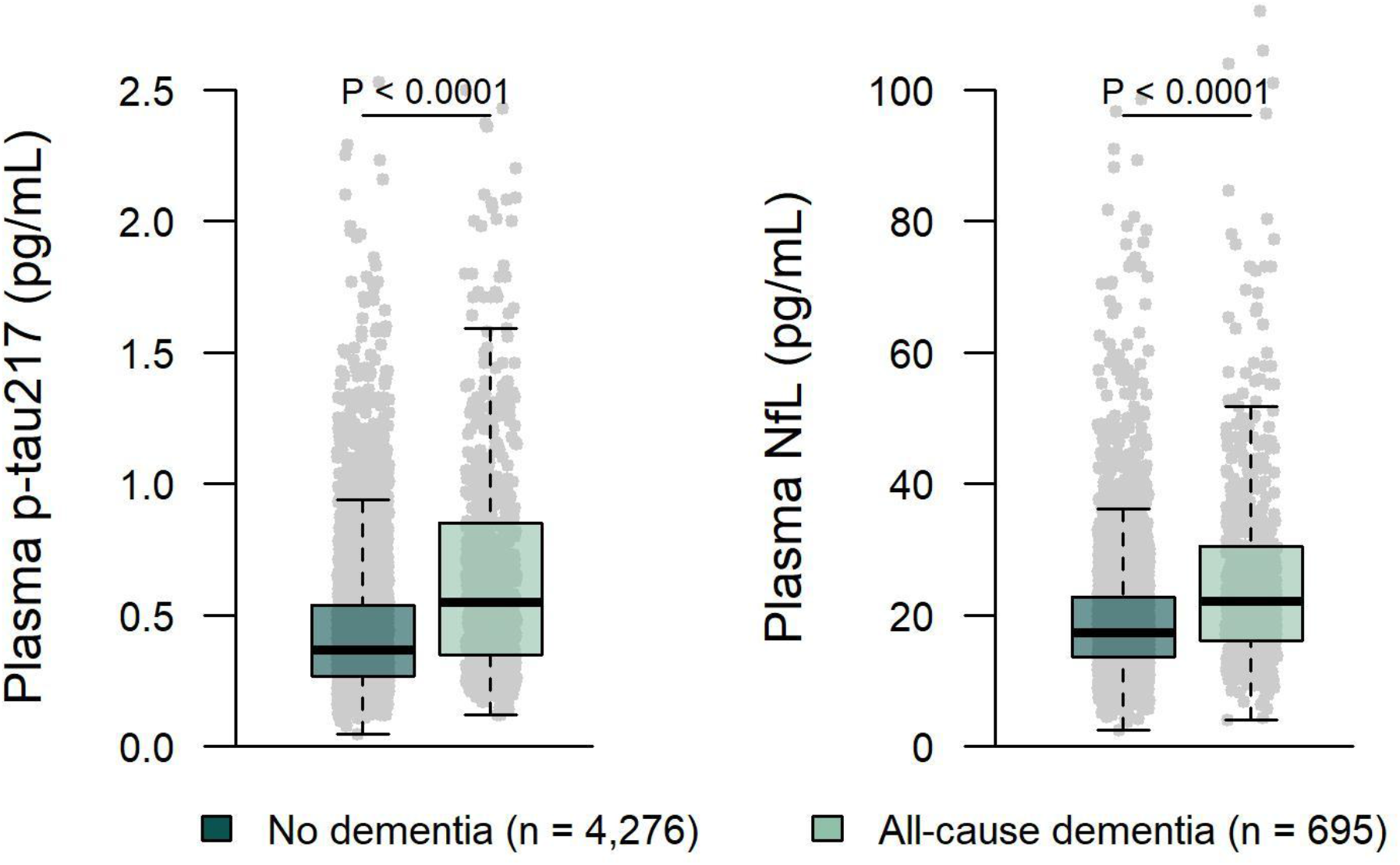
Baseline biomarker concentrations by incident all-cause dementia. Boxplots show the median (center line) and interquartile range (bounds of box) as well as the 2.5^th^ and 97.5^th^ percentiles (whiskers). Grey dots represent individual measurements. *P* values were derived from a two-sided Mann-Whitney test. p-tau217, phosphorylated tau at threonine 217; NfL, neurofilament light; pg/mL, picograms per milliliter.

### Risk estimations for incident dementia stratified by plasma p-tau217

The AR of developing dementia was 10.6% (95% CI 9.3-12.1) for participants in the low p-tau217 range and 33% (95% CI 29.9-36.1) for participants in the high range, with an AR difference of 22.5%, and a RR of 2.05 (95% CI 1.73–2.44). For the intermediate range, the AR was 16.9% (95% CI 14.6-19.5), with a RR of 1.23 (95% CI 1.01-1.50) (Tables S1 and S2, Figure 3). Assessing baseline cognitive status and p-tau217 together, AR was highest in the MCI group with high range p-tau217 (49.6%, 95% CI 44.7–54.5), and lowest in the CU group with low range p-tau217 (4.8%, 95% CI 3.8–6.0) (Table 2). Conversely, RR for incident dementia associated with elevated p-tau217 was higher within the CU group than within the MCI group (Table S2, Figure 3). In those with high p-tau217, females had a higher AR of dementia (37.7%, 95% CI 33.3-42.3) than males (27.6%, 95% CI 23.7-31.9) (Table S1), but after adjustment for age, education, *APOE* genotype, BMI and baseline cognition, RRs for females and males comparing high versus low p-tau217 range were not significantly different (Table S2).

**Figure 3.**
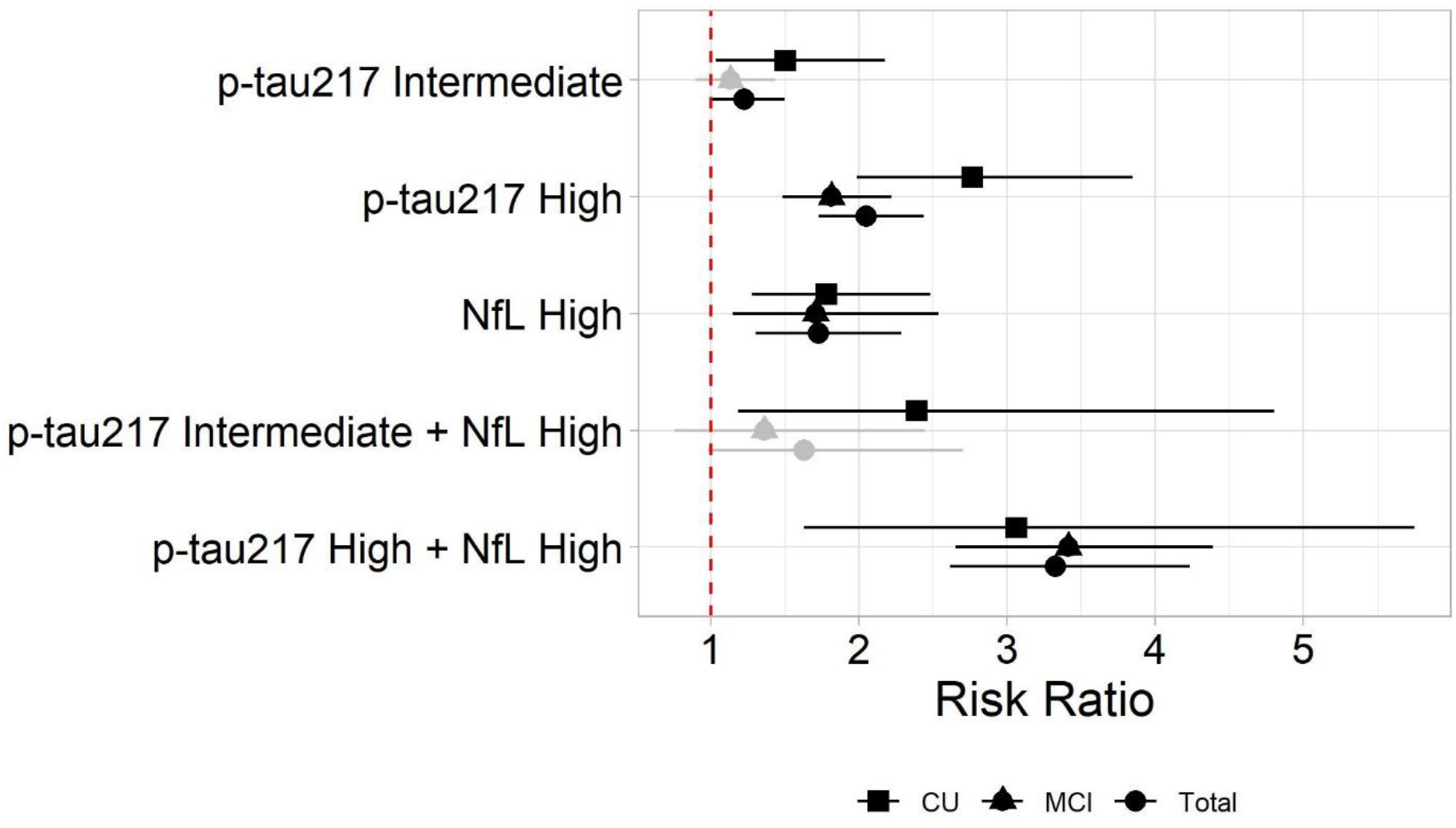
Risk ratios for incident all-cause dementia calculated relative to the low range for each biomarker. All estimates shown are weighted for participation bias. Whiskers represent 95% confidence intervals. The red striped lines marks RR 1. p-tau217, phosphorylated tau at threonine 217; NfL, neurofilament light; CU, cognitively unimpaired; MCI, mild cognitive impairment.

Those with high range p-tau217 had a higher AR than those with low range p-tau217 in age groups 70-84 years. The same tendency was observed for those 85 years and older but with wider confidence intervals (Table S3). RRs for high versus low p-tau217 were significant for all age groups up to 89 years. (Table S2, Figure 4).

**Figure 4.**
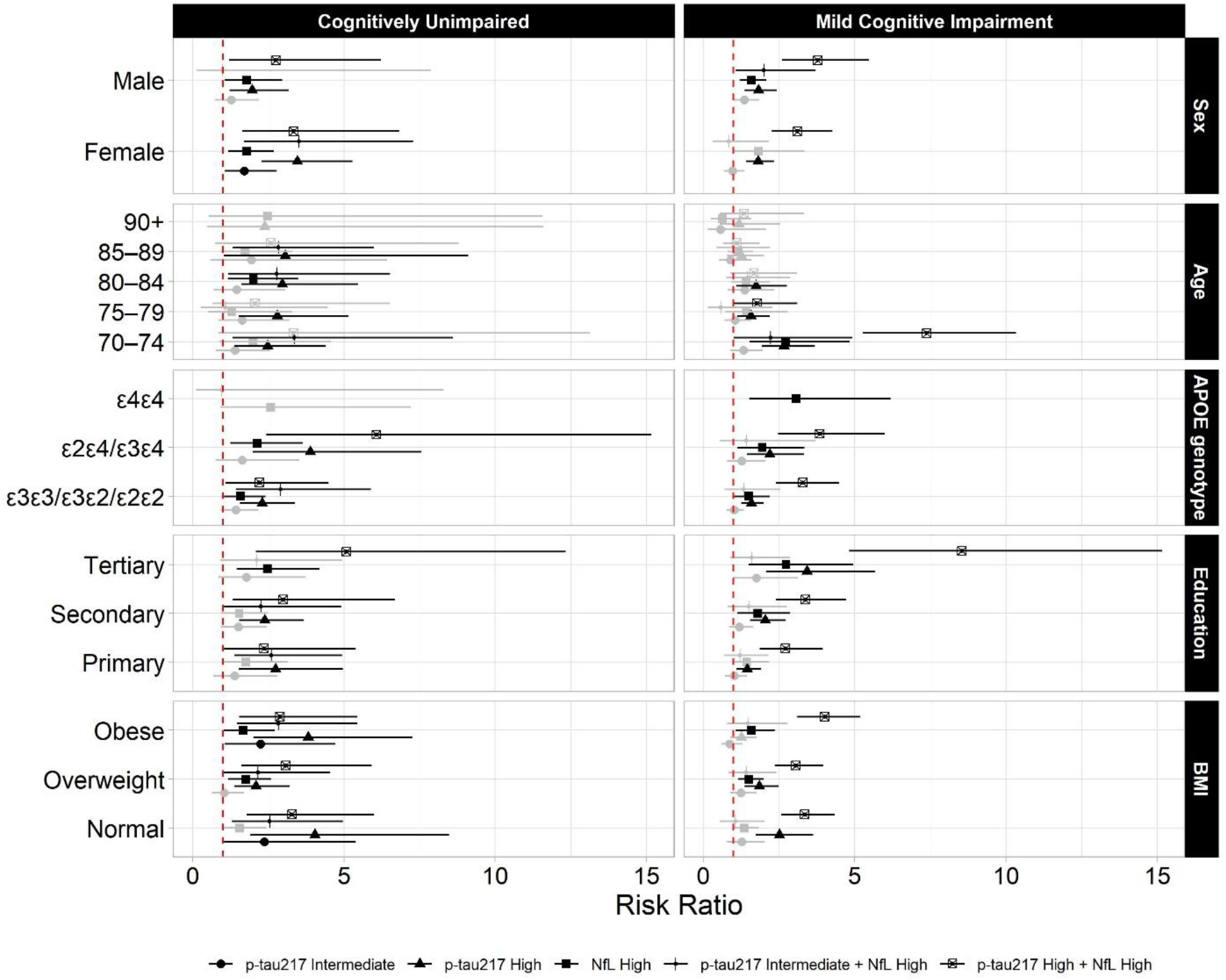
Risk ratios for all-cause dementia calculated relative to the low range for each biomarker. All estimates shown are weighted for participation bias. Whiskers represent 95% confidence intervals. Statistically significant risk ratios are shown in black. Non-significant risk ratios are shown in light grey. The red striped lines mark RR 1. p-tau217, phosphorylated tau at threonine 217; NfL, neurofilament light; *APOE*, apolipoprotein E; BMI, body mass index.

*APOE* ε4 carriers had higher dementia risk than non-carriers. Intermediate and high p-tau217 increased AR independent of *APOE* genotype. Among *APOE* ε4 heterozygotes and non-carriers, high range p-tau217, was associated with increased relative risk. Of those homozygous for *APOE* ε4 (N=109), only a small group had low range p-tau217 (n=14, 12.8%). None of these individuals developed dementia, and nearly all (n=12) were CU at baseline. With none of the low range p-tau217 *APOE* ε4 homozygous group developing dementia, it was not possible to model the RR for high/intermediate versus low p-tau217 range (Tables S2 and S4, Figure 4).

Lower education was associated with higher dementia risk. In all educational groups, the AR increased from low to intermediate to high p-tau217 ranges (Table S5). After adjustment for age, sex, *APOE* and BMI, high p-tau217 range was still associated with a higher RR in all educational and cognitive subgroup combinations (Table S2, Figure 4). There was no difference in dementia risk between BMI subgroups (Tables S2 and S6, Figure 4).

### Odds for incident dementia associated with continuous p-tau217 and NfL measurements

The predicted probability of all-cause incident dementia increased linearly with increasing p-tau217 concentrations. Sex-dependent differences in trajectories were seen, with ORs of 4.53 per 1 pg/mL increase in females and 1.54 per 1 pg/mL increase in males (*p* <.0001). Plasma NfL showed a significant non-linear association with incident dementia, with no differences between sexes. Baseline cognitive status did not modify the associations of p-tau217 or NfL with incident dementia, whereas the association between education and incident dementia differed between cognitively unimpaired and MCI participants. (Table S7, Figure 5).

**Figure 5.**
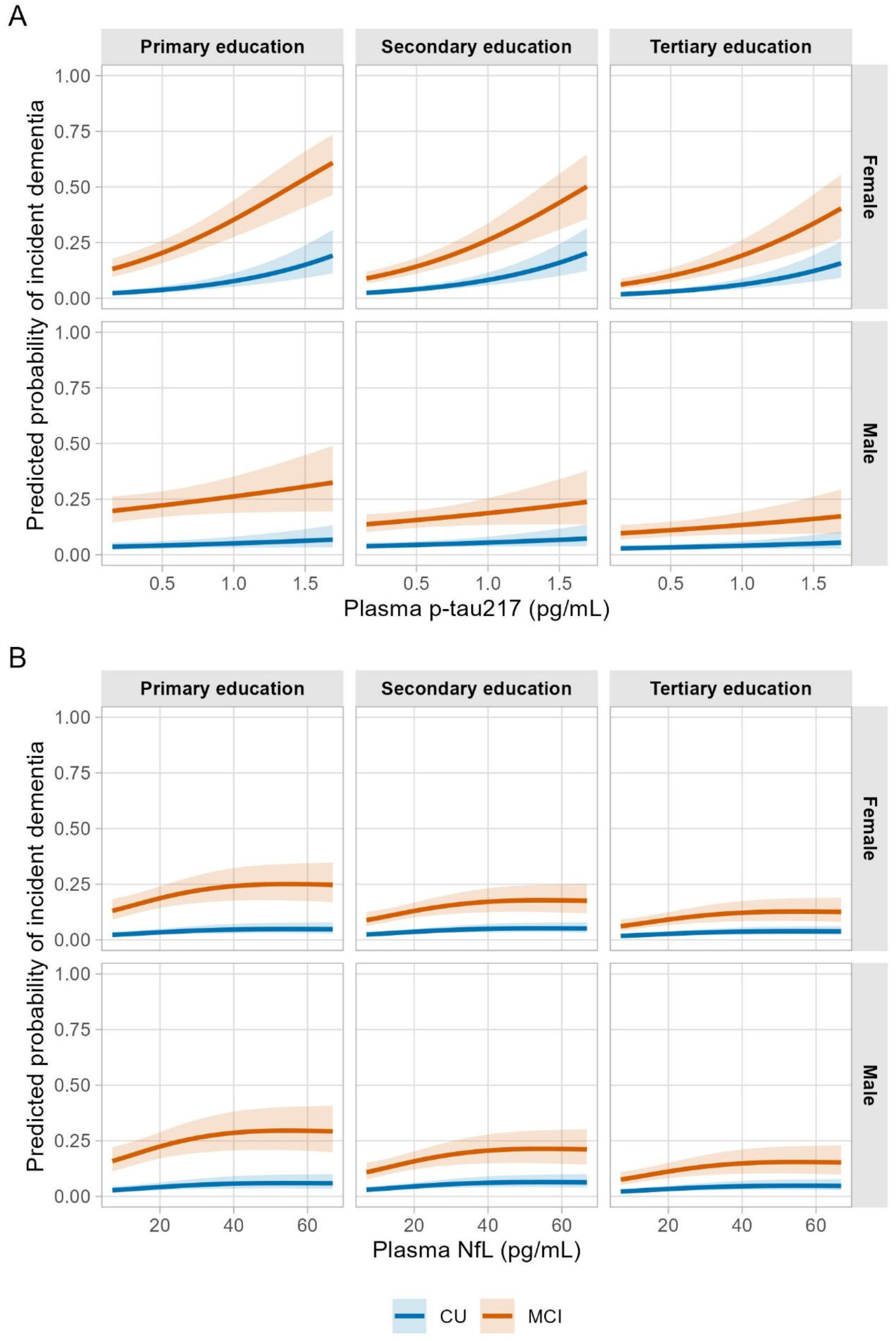
Graphical presentation of predicted probabilities for incident all-cause dementia associated with continuous plasma p-tau217 (Panel A) and NfL (Panel B) concentrations, adjusted for clinical covariates (cognitive status, sex, age group, apolipoprotein E genotype, serum creatinine, body mass index and educational level) from a Survey-Weighted Logistic Regression Model. CU, cognitively unimpaired; MCI, mild cognitive impairment; p-tau217, phosphorylated tau at threonine 217; NfL, neurofilament light; pg/mL, picograms per milliliter.

### Risk estimations for incident dementia stratified by plasma NfL

The AR of developing dementia was 15.7% (95% CI 14.4-17.0) for participants with low NfL range and 35.4% (95% CI 30.6-40.5) for participants with high range, with an AR difference of 19.7% and a RR of 1.72 (95% CI 1.31-2.27). Assessing baseline cognitive status and NfL range together, AR was highest in the MCI group with high plasma NfL (50.3%, 95% CI 42.8-57.8), and lowest in the CU group with low NfL (7.3%, 95% CI 6.3–8.5). There was no sex difference in dementia risk associated with high NfL (Tables S8-S10 and Figures 3 and 4).

In all *APOE* genotype subgroups, in all educational subgroups, and in all BMI subgroups, high NfL was associated with a higher dementia risk (Tables S9, S11-S13).

### Risk estimations for incident dementia stratified by combined plasma p-tau217 and NfL ranges

Participants with combined low p-tau217 and NfL had an AR of 10.5% (95% CI 9.1–12.0), while those with combined high biomarkers had an AR of 47.8% (95% CI 40.7–55.0), with an AR difference of 37.3% and a RR of 3.32 (95% CI 2.61-4.23) (Figure 3 and Tables S14 and S15). Combined high p-tau217 and NfL yielded a 3-fold RR in both the CU and MCI group. For those with MCI and biomarkers in the high range, AR for dementia was 64.2% (95% CI 53.8–73.5), while the CU group with low range biomarkers had an AR of 4.6% (95% CI 3.6–5.9). There were no significant risk differences between females and males (Tables S14-S15). Combined high p-tau217 and NfL was associated with increased risk for incident dementia in all age groups. Those aged 70-74 years, CU and in the low biomarkers range had a 3.1% risk of incident dementia (95% CI 2.1-4.5), while those aged 70-74 years, with MCI and high biomarkers had a 92.1% dementia risk (95% CI 70.9-98.2) (Tables S15-S16).

In the *APOE* ε4 heterozygous and non-carrier groups, high NfL contributed information on absolute dementia risk in addition to high p-tau217 (Tables S2, S4, S15, S17). Across educational and BMI subgroups, combined high p-tau217 and NfL were associated with increased dementia risk (Tables S2, S6, S15, S18-S19).

A forest plot for the multivariable-adjusted RR for incident dementia is shown in Figures 3 and 4.

Exploring baseline NfL biomarker status among participants with low baseline plasma p-tau217 (n = 2571) who had developed all-cause dementia at follow-up (n = 215), 199 had low NfL (92.6%; IPW 92.7%) and only 14 high NfL (6.5%; IPW 7.23%). Two participants were missing NfL data. Combined intermediate p-tau217 and high NfL was significantly associated with increased RR in groups with low AR: the cognitively unimpaired group (RR 2.39, 95% CI 1.19-4.81), age group 70-74 years (RR 2.52, 95% CI 1.18-5.38), *APOE* ε4 non-carriers (RR 1.74, 95% CI 1.02-2.98) and those with tertiary education (RR 1.78, 95% CI 1.01-3.15) (Table S20). Risk estimations for clinical ADD stratified by plasma p-tau217 and NfL mirror the results of the all-cause dementia analyses closely and can be found in the **Supplementary**.

### Influence of self-reported comorbidities on the association between biomarker range and incident dementia

Self-reported comorbidities, except for diabetes mellitus, did not significantly attenuate the RR for incident dementia associated with intermediate or high p-tau217, high NfL or concordant high p-tau217 and NfL, when adjusting for covariates. High p-tau217 range was less associated with incident dementia in those reporting diabetes compared to those that did not self-report diabetes, with a similar but non-significant trend observed in those reporting cardiovascular or cerebrovascular disease (Table S21).

## DISCUSSION

This largest-to-date population-based study with 4,971 dementia-free individuals aged 70 years and older found that plasma p-tau217 and NfL predicted 4-year dementia risk.

Our findings are in line with previous studies on more selected populations and a smaller population-based study, which show that plasma p-tau217 and NfL are associated with future cognitive decline in dementia-free individuals.[11, 13, 25, 26] For p-tau217, a double cut-off strategy was used to increase the overall accuracy for classifying individuals with or without Alzheimer’s pathology.[22] Previous analyses have estimated high range p-tau217 to have a positive and negative predictive value of 77.4% and 95.4% in the HUNT4 70+ population (optimism corrected 71.9% and 92.9%).[27] Plasma p-tau217 has recently gained broader implementation in clinical practice in those with MCI or dementia. However, there has been uncertainty about the clinical implication of intermediate range p-tau217. Our results showed that intermediate range p-tau217, and not only high range p-tau217, was associated with increased dementia risk. The probability of incident dementia was positively associated with p-tau217 concentration in a linear manner. This suggests that p-tau217 concentration provides relevant prognostic information beyond the predefined ranges for amyloid pathology. Plasma p-tau217 is considered a marker of amyloid pathology but concentrations increase further when Alzheimer’s related tauopathy develops and spreads.[21, 28, 29] Tauopathy is more strongly associated with cognitive impairment than amyloidosis.[30] Phosphorylated tau isoforms closer related to AD tauopathy than to amyloid pathology, like plasma microtubule-binding region-tau243, have been found to be closer associated with cognitive impairment than plasma p-tau217 and might be able to further refine risk estimates.[3, 31] However, as to date, no such immunoassays suitable for population-based studies are available.

We found no significant difference in relative risk associated with high range p-tau217 between sexes. This aligns with a recent prospective birth cohort study that found no association between sex and risk of incident dementia when adjusting for demographic factors and medical conditions,[32] and a systematic review and meta-analysis that found no sex differences in dementia incidence, except in the oldest-old.[33] Contrary, the predicted probability of dementia increased with increasing plasma p-tau217 concentrations above the upper threshold in a sex-specific manner. These findings may indicate that females are more susceptible to the adverse effects of advanced AD pathology than males. Previous research found that females with moderate/frequent amyloid plaques were more likely to reach high Braak tau stages than males[34] and that for a given concentration of plasma p-tau217, females showed greater cross-sectional and longitudinal tau accumulation across multiple brain regions and faster cognitive decline relative to males.[35] No sex differences for NfL and dementia risk were seen in our study.

Age is the most important risk factor for ADNC[27] and for dementia development.[36] Several studies have shown that p-tau217 is elevated in the presence of ADNC and not because of aging alone.[6, 21] In line with this, our data showed that high baseline p-tau217 was associated with an increased dementia risk in CU participants up to 89 years of age and in MCI participants up to 84 years of age. Mixed pathologies occur more common in the oldest-old and are known to be associated with a higher risk of dementia than just having “pure” AD.[37, 38] This might explain why high p-tau217 was less associated with incident dementia in the study’s CU participants ≥ 90 years and MCI participants ≥ 85 years, as these groups are expected to have more relevant co-pathologies than younger groups, which could weaken the association of p-tau217 with dementia as more “competing” co-pathologies are present. Another possibility is that these age groups contain too few participants to show statistically significant risk differences. Similarly, high NfL reaches statistical significance for the association with incident dementia in some but not all age groups, which could be due to small numbers. It has clinical implications that high NfL was found to be less common than intermediate and high p-tau217, as this implies that NfL, when used in unselected CU or MCI groups, would identify fewer at-dementia-risk people than p-tau217.

High p-tau217 was associated with higher dementia risk in all educational groups. In a recent HUNT publication, we showed that ADNC was more prevalent in those with low education.[27] Put together, the HUNT data indicate that higher education is associated with lower risk for development of ADNC, but once ADNC has been established, risk of developing dementia increases also in this group. In the present study, the risk of incident dementia in the low p-tau217 range differed significantly between the educational groups, with the highest risk in those with primary education. This could indicate that other factors than AD pathology contribute relatively more to dementia risk in low educated groups than in higher educated groups. Previous evidence suggests that increased dementia prevalence associated with low education in the HUNT cohort is partially mediated by various health and lifestyle factors from early to late adulthood, but that these factors explain less than one-fifth of the educational differences in dementia risk later in life.[39]

A limitation to our study is the lack of information on the cause of death of those deceased between baseline and follow-up. The otherwise rigorous application of IPW might attenuate this limitation. The HUNT study does not collect data on ethnicity, but the population in this region at the time of HUNT4 included less than 5% immigrants or Norwegian-born to immigrant parents from Africa, Asia, Middle-or South America. The data therefore represent a mainly Caucasian Norwegian population. The association between plasma p-tau217 and NfL and incident dementia could differ in other populations.[40]

We included body mass index in our analysis as there are data indicating that BMI and blood volume can influence the concentration of plasma p-tau217 and NfL, independent of brain amyloid burden, with high BMI being associated with “false-low” biomarker concentrations. This could potentially influence the predictive ability.[41] However, our data suggests that the ability of plasma p-tau217 and NfL to predict dementia incidence risk was independent of BMI. Our methodology is not suitable to address causality between BMI and dementia incidence. Our data showed that a high p-tau217 range was less associated with incident dementia in those reporting diabetes compared to those that did not self-report diabetes, with a similar but non-significant trend observed in those reporting cardiovascular or cerebrovascular disease. We tested our model both with and without serum creatinine as a covariate to differ if this might be caused by a higher prevalence of impaired kidney function in these groups, which could cause “false-high” p-tau217 concentrations. Adding serum creatinine to the model did not influence this finding. Systematic reviews and meta-analysis have shown some glucose-lowering therapies to be associated with reduced dementia risk,[42, 43] and further studies should explore this possible association in the HUNT population.

A limitation to our study is that dementia incidence was assessed only once, four years after baseline. Higher plasma p-tau217 and NfL concentrations have been shown to be associated with faster progression from MCI to all-cause and AD dementia in a study of up to 16 years of follow-up.[44] It is also a limitation that the diagnosis of dementia subtypes was based entirely on clinical information, without biomarker data to support etiological considerations, but this has no influence on the results of the all-cause dementia analysis. Of those potentially eligible for follow-up, 9% had deceased. It is a limitation that we do not have information on their cause of death, but the otherwise rigorous application of IPW might attenuate this limitation. The HUNT study does not collect data on ethnicity, but the population in this region at the time of HUNT4 included less than 5% immigrants or Norwegian-born to immigrant parents from Africa, Asia, Middle-or South America. The data therefore represent a mainly Caucasian Norwegian population. The association between plasma p-tau217 and NfL with incident dementia could differ in other populations.[40]

The biggest strength of the study is the populations-based design, the inclusion of nearly 5,000 participants, the HUNT study population‘s close alignment with key Norwegian demographic and health indicators, the high participation rate from baseline to follow-up and the use of extended inverse probability weighting to adjust for selection bias. Home visits and inclusion of those living in nursing homes further mitigated inclusion bias. Further strengths are the use of a standardized, thorough cognitive assessment and the use of externally validated biomarker cut-offs.

As of today, it is not recommended to screen cognitively unimpaired people for ADNC outside of research settings.[45] Screening recommendations could change if effective, safe and cost-efficient disease-modifying treatment for preclinical AD should become available. For those with prodromal AD, amyloid targeting therapies are already available in some countries. The incentive to test people with MCI for ADNC will largely depend on whether specific treatment options are available or not. The possible psychological impact of receiving a high-risk result should be discussed before administering prognostic testing.[46]

In conclusion, these Norwegian population-based data show that plasma p-tau217 and NfL independently and jointly predict 4-year incident dementia risk in people aged ≥70 years.

**Table 1.** Baseline characteristics of study participants overall and by incident all-cause dementia at follow-up.

| Characteristics<br>at baseline | Diagnosis at follow-up |  | Total | P-values |
| --- | --- | --- | --- | --- |
|  | No dementia | Dementia |  |  |
| Total, n (%) | 4,276 (86.0) | 695 (14.0) | 4,971 (100.0) |  |
| Age, (years, mean ± s.d.) | 75.4 ± 4.31 | 78.6 ± 5.90 | 75.8 ± 4.69 | <.0001 |
| Female, n (%) | 2,264 (52.9) | 378 (54.4) | 2,642 (53.1) | .4863 |
| Education, n (%) |  |  |  |  |
| *Primary* | 894 (20.9) | 233 (33.5) | 1,127 (22.7) | <.0001 |
| *Secondary* | 1,968 (46.0) | 322 (46.3) | 2,290 (46.1) |  |
| *Tertiary* | 1,412 (33.0) | 138 (19.9) | 1,550 (31.2) |  |
| *Missing* | 2 (0.05) | 2 (0.29) | 4 (0.08) |  |
| MoCA, (mean ± s.d.) | 24.8 ± 2.78 | 21.6 ± 3.10 | 24.4 ± 3.03 | <.0001 |
| *APOE* genotype |  |  |  |  |
| $\epsilon 3\epsilon 3/\epsilon 3\epsilon 2/\epsilon 2\epsilon 2$ | 3,078 (72.0) | 442 (63.6) | 3,520 (70.8) | <.0001 |
| $\epsilon 2\epsilon 4/\epsilon 3\epsilon 4$ | 1,100 (25.7) | 215 (30.9) | 1,315 (26.5) | |
| $\epsilon 4\epsilon 4$ | 75 (1.75) | 34 (4.89) | 109 (2.19) | |
| *Missing* | 23 (0.54) | 4 (0.58) | 27 (0.54) |  |
| Clinical diagnosis, n (%) |  |  |  | <.0001 |
| *CU* | 2,937 (68.7) | 233 (33.5) | 3,170 (63.8) |  |
| *MCI* | 1,338 (31.3) | 461 (66.3) | 1,799 (36.2) |  |
| Plasma p-tau217 pg/mL, (mean ± s.d.) | 0.45 ± 0.32 | 0.67 ± 0.46 | 0.48 ± 0.35 | <.0001 |
| Plasma NfL pg/mL, (mean ± s.d.) | 19.8 ± 13.3 | 26.3 ± 18.7 | 20.7 ± 14.3 | <.0001 |
| BMI (mean ± s.d.) | 27.3 ± 4.17 | 27.2 ± 4.35 | 27.3 ± 4.20 | .6086 |

s.d., standard deviation; MoCA, Montreal Cognitive Assessment; *APOE*, apolipoprotein E; p-tau217, phosphorylated tau at threonine 217; NfL, neurofilament light; pg/mL, picograms per milliliter; BMI, body mass index. ‘No dementia’ included participants with normal cognition, mild cognitive impairment, and those defined as ‘other cognitive impairment’, meaning those with cognitive impairment who did not fulfill DSM-5 criteria for minor or major neurocognitive disorder.

## AUTHOR CONTRIBUTIONS

Conceptualization: ALS, DTR, HKS, GS, NJA, DA

Methodology: ALS, DTR, GS, NJA, DA

Coordinated blood sample collection and quality assurance: MN

Coordinated and/or performed blood biomarker quantification: KT, BA, HH, IP, GDM, HZ, NJA

Data analysis: DTR

Writing - Original Draft: ALS, DA, DTR, GS, HKS, NJA

Writing - Review & Editing: ALS, DA, DTR, GS, HKS, NJA, AOVM, KP, KT, BA, HH, IP, GDM, HZ, MN

Visualization: DTR

Project administration: ALS, DA

Funding acquisition: ALS, DTR, HZ, HKS, GS, NJA, DA

All authors have reviewed and agreed to the final manuscript draft.

## CONSENT STATEMENT

This study was approved by the Regional Committee for Medical and Health Research Ethics in Norway (REC Southeast C 565876). Participation in HUNT required informed consent. In participants with reduced capacity to consent, next of kin gave consent. Data were collected in accordance with the Declaration of Helsinki.

## Data Availability

To protect participants privacy, HUNT Research Centre aims to limit storage of data outside HUNT databank and cannot deposit data in open repositories. HUNT databank has precise information on all data exported to different projects and can reproduce them on request. There are no restrictions regarding data export given approval of applications to HUNT Research Centre. Researchers can apply for data at https://www.ntnu.edu/hunt/research

