## Supplementary for "Plasma p-tau217 and NfL predict incident dementia in the community"

This appendix has been provided by the authors to give readers additional information about their work.

### Supplementary methods

#### Cognitive assessment and diagnosis

In both HUNT4 70+ and HUNT AiT, trained health personnel assessed the participants' cognitive, neuropsychiatric, and functional status using standardized interviews and clinical assessments at field stations, at homes, or in nursing homes as previously described.[1-3] Cognition was assessed with the Norwegian version of the Montreal Cognitive Assessment (MoCA). The Word List Memory Task (WLMT) from the Consortium to Establish a Registry of Alzheimer's disease (CERAD)[4] was additionally conducted in those participants scoring ≥ 22 points on the MoCA and remembering at least 1 of the 5 words on the subtask on delayed recall in MoCA. Participants also completed the Meta-Memory Questionnaire, a nine-item self-report questionnaire designed to assess subjective memory performance.[5] Participants were interviewed about subjective cognitive impairments, the development pattern of cognitive impairment, function in activities of daily living, neuropsychiatric symptoms, former examinations on cognitive function, and family history of cognitive impairments. When cognitive impairment was indicated, consent was obtained for a structured interview with a proxy or, in nursing homes, a professional caregiver, to gather information on symptom onset, disease course, neuropsychiatric symptoms (assessed with the 12-item Neuropsychiatric Inventory Questionnaire),[6] physical activities of daily living[7] and instrumental activities of daily living. For participants living in nursing homes, the Severe Impairment Battery-8[8] was used instead of MoCA when the structured interview with the proxy or professional caregiver indicated moderate to severe dementia. The overall level of cognitive and functional impairment was assessed with the Clinical Dementia Rating Scale (CDR).[9] The depression subscale from The Hospital Anxiety and Depression Scale was used to screen for depression.[10] The Short Physical Performance Battery (SPPB) was used to test physical function.[11]

For each participant, two out of a pool of several (nine at HUNT4 70+, eight at AiT) research physicians with clinical expertise (geriatrics, neurology, old-age psychiatry) made independent diagnoses according to

The Diagnostic and Statistical Manual of Mental Disorders, Fifth Edition (DSM-5) criteria (cognitively unimpaired (CU), mild cognitive impairment (MCI) (minor neurocognitive disorder in DSM-5) or dementia (major neurocognitive disorder in DSM-5).[12] In cases where consensus was not reached, a third physician was consulted. People with a diagnosis of dementia were sub-classified as having clinical Alzheimer's disease dementia, vascular dementia, Lewy body dementia (including dementia with Lewy bodies and Parkinson's disease dementia), frontotemporal dementia, mixed dementia, other specified dementia, or unspecified dementia based on all the available information. Mixed dementia was defined as dementia due to multiple etiologies in line with the DSM-5 criteria for Major Neurocognitive Disorder due to Multiple Etiologies. The detailed diagnostic procedure has been published previously.[1-3] Biomarker data were not available when the clinical diagnoses were made, nor were brain imaging data. Research physicians assessing the cognitive status at HUNT AiT were blinded to the previous cognitive diagnosis from HUNT4 70+.

A small number of participants (181 participants in HUNT4 70+ and 64 in the nested HUNT AiT dataset) did not have sufficient information to be classified according to the DSM-5 criteria and were excluded. This was due to missing information – either refusal to undergo cognitive testing or lack of available proxy information. An even smaller number of participants (5 each in HUNT4 70+ and AiT) were assessed to have cognitive impairment other than DSM-5 classified minor or major neurocognitive disorder.

##### *APOE* genotype, comorbidities, body mass index and education levels

*APOE* genotype data were readily available from the HUNT biobank.[3] Comorbidities and education levels were self-reported, using standardized questionnaires. The presence of comorbidities was assessed by yes/no questions ("Have you had, or do you have any of the following diseases?"). For this study, to increase statistical power, "angina pectoris" and "myocardial infarction" were merged as "cardiovascular disease". "Diabetes" and "Has it ever been verified that you had high blood sugar (hyperglycaemia)?" was merged into the category "diabetes mellitus". "Rheumatoid arthritis" and "spondyloarthritis" were merged into one category. "Stroke/brain haemorrhage" was not merged with any other comorbidity class, neither was "COPD or emphysema". Education was defined as primary, secondary and tertiary education. "Primary education" included up to 10 years of compulsory primary and lower secondary education; "Secondary education" was defined as additional 1–2 years of academic or vocational school, 3 years of academic or vocational school, or 3–4 years of vocational training or apprenticeship (upper secondary education); "Tertiary education" referred to college or university education. Body mass index (BMI, kg/m²) was measured by the InBody770 and categorized into underweight (<18.5 kg/m²), normal weight (18.5-24.9 kg/m²), overweight (25-29.9 kg/m²) and obesity (≥30 kg/m²).

#### Blood sample collection and handling procedures

Blood samples were collected, handled, and stored according to standardized procedures.[13] Samples were either collected at field stations, in participants' homes or in nursing homes. Fasting was not required. Samples were collected in Vacuette® Ethylene Diamine Tetra acetic Acid (EDTA)-plasma tubes 9ml that were gently inverted 6-8 times after sampling. After a maximum of 45 to 120 minutes at room temperature at HUNT4 and after 15 minutes at HUNT AiT, samples were placed in a refrigerator with a temperature of 2-8 °C before transportation at 4 °C to a central laboratory the same day. The next day, plasma samples were centrifuged at 2500 x g for 15 minutes at 6 °C, before undergoing automated fractioning (Tecan200). Plasma aliquots (tube size 1.4 mL containing 200μl plasma) were frozen to -80 °C and stored until transportation for future analyses. Thus, phlebotomy-to-freezing time ensured plasma p-tau217 and NfL stability.[14] Transportation from the HUNT biobank to the Clinical Neurochemistry Laboratory, University of Gothenburg (Sweden) was conducted with temperature-regulated dry-ice transport at -80 °C. Samples were further stored at -80 °C until analysis. Prior to immunoassay analyses, plasma-EDTA samples were thawed, vortexed and centrifuged at 4000 x g for 10 minutes at 20 ºC.

#### Analysis of plasma p-tau217 and NfL

Biomarkers were analyzed on the Simoa HD-X instrument at the University of Gothenburg, Sweden, according to the manufacturer's instructions (Quanterix, Billerica, MA, USA). Plasma p-tau217 concentration was measured with a previously validated commercial kit (ALZpath p-Tau 217 Advantage PLUS, Quanterix).[15] As recommended by the Global CEO Initiative on AD, we used a two cut-off approach to categorize individuals as ADNC negative, intermediate, or positive, using validated plasma p-tau217 cut-offs: lower cut-off 0.40 picograms per milliliter (pg/mL, 95% sensitivity) and an upper cut-off of ≥ 0.63 pg/mL (95% specificity).[15, 16] We used the upper cut-off to determine with a high specificity those with the presence of ADNC, and the lower cut-off to identify those with a high likelihood of not having ADNC, with an intermediate range between these cut-offs. Plasma NfL concentration was measured using a commercially available immunoassay from Quanterix (Simoa® NF-light Kit). Age-dependent NfL cut-offs were used, as previously recommended.[17] For age 70 years, we administered a cut-off of 20 pg/mL, and for age group ≥ 71 years a cut-off of 35 pg/mL, with concentrations above the cut-off indicating the presence of neuroaxonal damage.[18] Plasma analysis was conducted January to August 2024.

#### Summary of the analytical performance – Quanterix ALZpath p-Tau-217 Advantage PLUS

Calibrators were run in duplicate, and obvious outlier replicates were excluded prior to curve fitting.
Three quality control (QC) levels (human pooled plasma samples) were run in duplicate at the beginning and end of each run. Repeatability (% CVr); Intermediate precision (%CVRw)
At a concentration of 0.4 pg/mL (mean value), the repeatability was 10.3%, and the intermediate precision was 10.3%. At a concentration of 1.5 pg/mL (mean value), the repeatability was 10.6%, and the intermediate precision was 15.9%. At a concentration of 1.9 pg/mL (mean value), the repeatability was 8.9%, and the intermediate precision was 15.8%.

#### Summary of the analytical performance – Quanterix NfL Advantage V2 Kit

Two QC levels (human pooled plasma samples) were run in duplicate at the beginning and end of each run.
At a concentration of 14.2 pg/ml (mean value), the repeatability was 12.9 % and the intermediate precision

was 12.9 %. At a concentration of 89.8 pg/mL, the repeatability was 15.7 % and the intermediate precision was 15.7%

#### Statistical analysis

To account for differential participation and potential selection bias across study phases, inverse probability weighting was applied. Participation probabilities were estimated sequentially using logistic regression models. First, age-, sex-, and education-specific probabilities of participation in HUNT4 70+ were derived from published participation estimates and inverted to obtain initial weights.[19] Second, the probability of providing a blood sample at HUNT4 70+ was modelled as a function of cognitive status, age group, sex, education, and *APOE* ε4 genotype. Third, participation in the HUNT AiT sub-study was modelled using logistic regression adjusting for baseline (HUNT4 70+) cognitive status, age group, sex, education, *APOE* genotype, depressive symptoms (HADS), global cognitive performance (MoCA), and physical function (SPPB). Final analytic weights were defined as the inverse product of these probabilities. To improve numerical stability, extreme weights (n=9) were truncated at three interquartile ranges from the median.

All inferential analyses incorporated these inverse probability weights. Dementia outcomes included all-cause dementia and clinically diagnosed Alzheimer's disease dementia at the HUNT AiT assessment. Plasma p-tau217 concentrations were categorized as low (<0.40 pg/mL), intermediate (0.40–0.62 pg/mL), or high (≥0.63 pg/mL) range. Plasma NfL was dichotomized into low or high range using age-specific thresholds (<20 pg/mL for participants younger than 71 years and <35 pg/mL for participants ≥ 71 years). Combined biomarker variables were defined based on concordant high p-tau217 and high NfL range, and by concordant intermediate p-tau217 and high NfL range.

Weighted cumulative absolute risk (AR) and risk ratios (RRs) for dementia were estimated using inverse probability-weighted proportions with corresponding 95% confidence intervals, stratified by baseline cognitive status (CU vs MCI), sex, age group, *APOE* genotype, BMI and educational level. Associations between biomarker ranges and dementia outcomes were quantified using inverse probability-weighted generalized linear models with a quasi-Poisson distribution and log link to estimate RRs and 95% confidence intervals. Models included interaction terms between biomarker categories and baseline cognitive status, and stratified analyses were conducted to evaluate effect modification by demographic and genetic factors. We explored the inclusion of time between baseline and follow-up in the models without finding a significant contribution. Pairwise contrasts between biomarker categories were estimated on the log scale and exponentiated to derive RRs. The presence of significant differences in RR between biomarker ranges across sex was evaluated by rate of risk ratios tests.

Odds ratios (OR) for all-cause dementia associated with continuous plasma p-tau217 concentrations, NfL, and clinical covariates (cognitive status, sex, age group, serum creatinine, *APOE* genotype, BMI and educational level) were explored with a Survey-Weighted Logistic Regression Model. Continuous plasma p-tau217 was included linearly, and NfL with a second-order spline.

Additional analyses assessed whether associations between plasma p-tau217 range, NfL range and dementia risk differed according to major baseline comorbidities, including cerebrovascular disease, chronic obstructive pulmonary disease, cardiovascular disease, diabetes mellitus, and inflammatory rheumatic conditions. These models adjusted for age, sex, education, *APOE* genotype, serum creatinine and baseline cognitive status, and included interaction terms between biomarker ranges and each comorbidity.

Distributions of plasma p-tau217 and NfL according to dementia outcomes were compared using Wilcoxon rank-sum tests and visualized using boxplots with overlaid individual observations. All statistical analyses were performed using R 4.5.2. Statistical tests were two-sided, and a $P$ value $<0.05$ was considered statistically significant.

#### Supplementary results

Clinically diagnosed Alzheimer's disease dementia (furthermore abbreviated to ADD) at follow-up (HUNT AiT) was defined as a supplementary outcome. Median baseline concentration of plasma p-tau217 and NfL was higher in those who developed incident ADD compared to those who did not (Figure S1).

##### Risk estimations for incident ADD stratified by plasma p-tau217

Risk estimations for clinical ADD stratified by baseline plasma p-tau217 mirror the results of the all-cause dementia analyses closely, considering that a smaller proportion of the population received an ADD diagnosis than an all-cause dementia diagnosis. Details can be found in Tables S22-S27 and Figure S2.

##### Risk estimations for incident ADD stratified by plasma NfL

Risk estimations for clinical ADD stratified by baseline plasma NfL closely aligned to the results from the all-cause dementia analyses. Considering that a smaller proportion of the population received an ADD diagnosis than an all-cause dementia diagnosis, fewer sub-group results were statistically significant. Details can be found in Tables S28-S33 and Figure S2.

##### Risk estimations for incident ADD stratified by combining high plasma p-tau217 and NfL

Risk estimations for clinical ADD stratified by concordant high p-tau217 and NfL were similar to the estimations for the all-cause dementia analyses, when considering that a smaller proportion of the population received an ADD diagnosis than an all-cause dementia diagnosis. Contrary to the all-cause dementia analysis, females with MCI and high p-tau217 and NfL range had a higher absolute risk for ADD than males (Tables S11 and S32), while the RR was not significantly different. More details can be found in Tables S34-S39 and Figure S2.

#### Supplementary figures

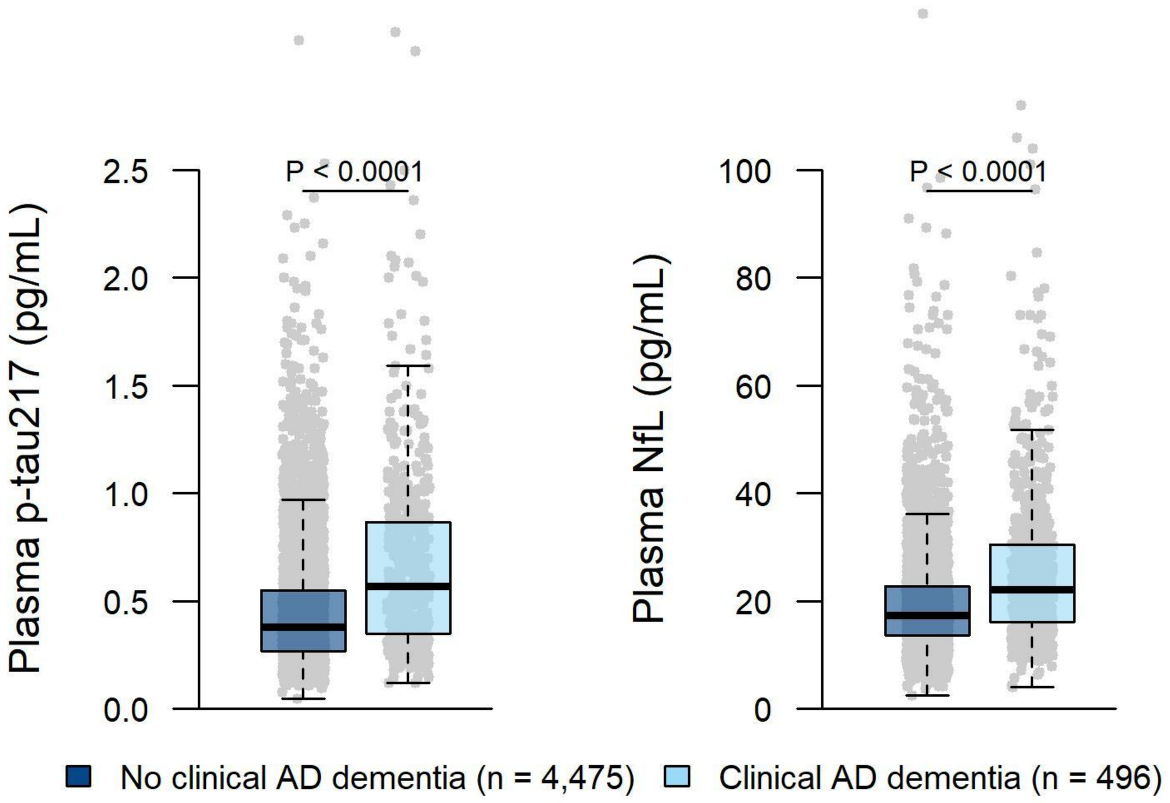

**Figure S1.** Baseline biomarker concentrations by incident Alzheimer's disease dementia. Boxplots show the median (center line) and interquartile range (bounds of box) as well as the $2.5^{th}$ and $97.5^{th}$ percentiles (whiskers). Grey dots represent individual measurements. *P* values were derived from a two-sided Mann-Whitney test. The group labeled 'No clinical AD dementia' contained cognitively unimpaired participants, participants with mild cognitive impairment, other cognitive impairment and dementia of other sub-types than Alzheimer's disease (vascular dementia, dementia with Lewy bodies, Parkinson's disease dementia, frontotemporal dementia, mixed dementia, other specified dementia, or unspecified dementia) at follow-up. Biomarker data were not available when the clinical diagnoses were made, nor were brain imaging data. p-tau217, phosphorylated tau at threonine 217; NfL, neurofilament light; pg/mL=picograms per milliliter.

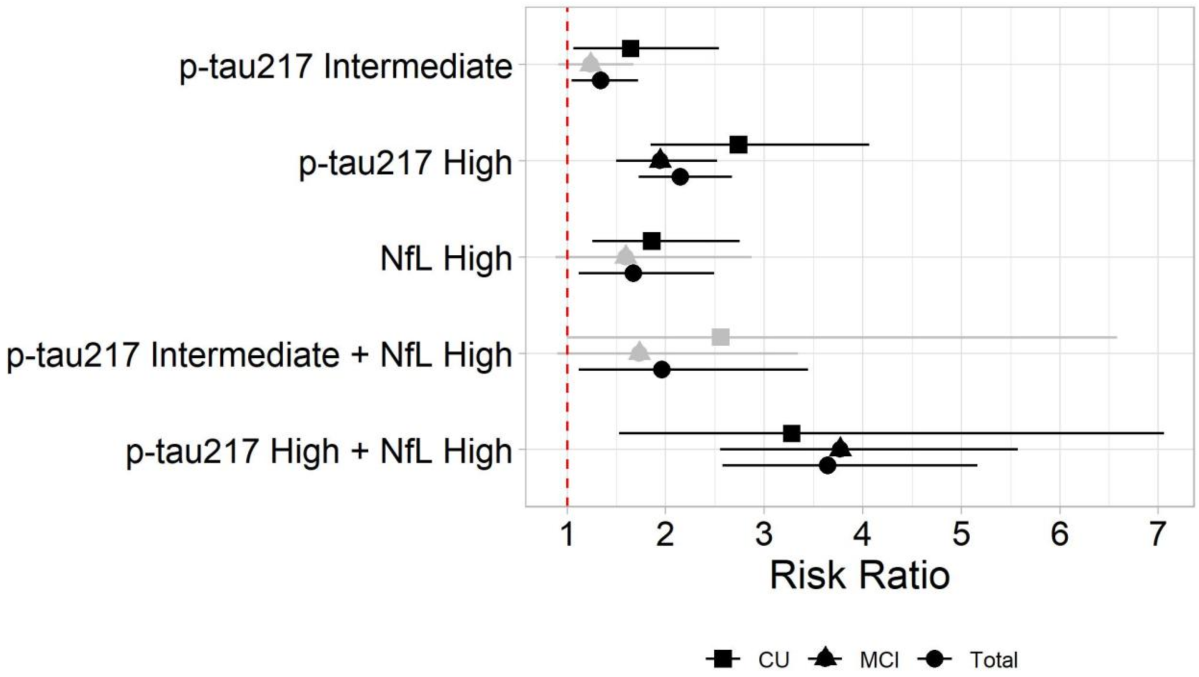

**Figure S2.** Risk ratios for incident Alzheimer's disease dementia are calculated relative to the low range for each biomarker. All estimates shown are weighted for participation bias. Whiskers represent 95% confidence intervals. The red striped lines marks RR 1. p-tau217, phosphorylated tau at threonine 217; NfL, neurofilament light; CU, cognitively unimpaired; MCI, mild cognitive impairment.

### Supplementary tables

#### Supplementary tables on p-tau217 and all-cause dementia

Table S1. Estimated absolute risk of all-cause dementia by baseline cognitive status, plasma p-tau217 range and sex.

| Clinical Diagnosis at baseline | Plasma p-tau217 at baseline | Female | | | Male | | | Total | | |
|---|---|---|---|---|---|---|---|---|---|---|
| | | N | Risk (95% CI) | | N | Risk (95% CI) | | N | Risk (95% CI) | |
| CU | < 0.40 | 995 | 4.9 | (3.6–6.6) | 721 | 4.6 | (3.2–6.6) | 1716 | 4.8 | (3.8–6.0) |
| | 0.40–0.63 | 428 | 9.0 | (6.3–12.6) | 383 | 7.5 | (5.1–10.9) | 811 | 8.3 | (6.4–10.7) |
| | ≥ 0.63 | 323 | 20.7 | (16.2–25.9) | 320 | 14.7 | (10.9–19.5) | 643 | 17.8 | (14.8–21.3) |
| | All | 1746 | 9.0 | (7.6–10.7) | 1424 | 7.8 | (6.4–9.5) | 3170 | 8.5 | (7.5–9.6) |
| MCI | < 0.40 | 435 | 22.8 | (18.5–27.7) | 420 | 17.6 | (14.0–21.9) | 855 | 20.3 | (17.4–23.6) |
| | 0.40–0.63 | 229 | 28.6 | (22.3–35.9) | 258 | 29.1 | (23.2–35.8) | 487 | 28.9 | (24.4–33.7) |
| | ≥ 0.63 | 230 | 55.4 | (48.5–62.1) | 227 | 42.6 | (36.0–49.6) | 457 | 49.6 | (44.7–54.5) |
| | All | 894 | 33.8 | (30.4–37.5) | 905 | 27.6 | (24.5–31.0) | 1799 | 30.9 | (28.5–33.3) |
| Total | < 0.40 | 1430 | 11.1 | (9.3–13.2) | 1141 | 9.9 | (8.1–12.0) | 2571 | 10.6 | (9.3–12.1) |
| | 0.40–0.63 | 657 | 16.8 | (13.6–20.5) | 641 | 17.1 | (14.0–20.8) | 1298 | 16.9 | (14.6–19.5) |
| | ≥ 0.63 | 554 | 37.7 | (33.3–42.3) | 547 | 27.6 | (23.7–31.9) | 1101 | 33.0 | (29.9–36.1) |
| | All | 2642 | 18.8 | (17.1–20.7) | 2329 | 16.4 | (14.7–18.2) | 4971 | 17.7 | (16.5–19.0) |

p-tau217, phosphorylated tau at threonine 217; CI, confidence interval; CU, cognitively unimpaired; MCI, mild cognitive impairment. Concentration is given in picograms per milliliter. Plasma p-tau217 data was missing from one participant. Two participants had cognitive impairment other than MCI. Estimates are weighted.

Table S2. Association between plasma p-tau217 range at HUNT4 70+ and risk of all-cause dementia at follow-up across demographic and clinical subgroups (N, 4,857).

| Characteristics at baseline | CU | MCI | Total | CU | MCI | Total |
|---|---|---|---|---|---|---|
| | RR (95% CI) | RR (95% CI) | RR (95% CI) | RR (95% CI) | RR (95% CI) | RR (95% CI) |
| | *Intermediate range plasma p-tau217* | | | *High range plasma p-tau217* | | |
| **All** | 1.50 (1.04–2.18) | 1.14 (0.90–1.44) | 1.23 (1.01–1.50) | 2.77 (1.99–3.85) | 1.82 (1.49–2.22) | 2.05 (1.73–2.44) |
| **Sex** | | | | | | |
| *Female* | 1.71 (1.05–2.78) | 0.96 (0.68–1.36) | 1.14 (0.86–1.51) | 3.46 (2.27–5.28) | 1.81 (1.41–2.33) | 2.21 (1.78–2.75) |
| *Male* | 1.27 (0.74–2.18) | 1.36 (1.00–1.85) | 1.33 (1.02–1.75) | 1.97 (1.22–3.19) | 1.82 (1.36–2.43) | 1.86 (1.45–2.38) |
| **Age** | | | | | | |
| *70–74* | 1.40 (0.76–2.58) | 1.32 (0.88–1.98) | 1.34 (0.96–1.88) | 2.48 (1.39–4.41) | 2.67 (1.94–3.68) | 2.62 (1.98–3.47) |
| *75–79* | 1.64 (0.84–3.19) | 1.05 (0.70–1.57) | 1.18 (0.84–1.66) | 2.80 (1.52–5.16) | 1.58 (1.13–2.20) | 1.84 (1.38–2.47) |
| *80–84* | 1.46 (0.70–3.06) | 1.37 (0.81–2.33) | 1.40 (0.91–2.16) | 2.97 (1.61–5.48) | 1.73 (1.09–2.77) | 2.12 (1.46–3.09) |
| *85–89* | 1.94 (0.59–6.43) | 0.90 (0.51–1.59) | 1.09 (0.65–1.82) | 3.05 (1.02–9.11) | 1.26 (0.79–2.00) | 1.58 (1.03–2.45) |
| *90+* | - - | 0.57 (0.16–2.07) | 0.44 (0.12–1.54) | 2.39 (0.49–11.6) | 1.17 (0.54–2.54) | 1.45 (0.71–2.95) |
| ***APOE* genotype** | | | | | | |
| *ε3ε3/ε3ε2/ε2ε2* | 1.43 (0.93–2.18) | 1.02 (0.78–1.34) | 1.13 (0.90–1.41) | 2.29 (1.56–3.37) | 1.58 (1.26–1.99) | 1.76 (1.45–2.15) |
| *ε2ε4/ε3ε4* | 1.64 (0.76–3.52) | 1.27 (0.79–2.06) | 1.36 (0.90–2.04) | 3.88 (1.99–7.56) | 2.20 (1.45–3.33) | 2.58 (1.80–3.69) |
| *ε4ε4* | - - | - - | - - | - - | - - | - - |
| **Education** | | | | | | |
| *Primary* | 1.38 (0.68–2.81) | 1.02 (0.72–1.44) | 1.08 (0.80–1.48) | 2.75 (1.52–4.96) | 1.45 (1.10–1.91) | 1.68 (1.30–2.15) |
| *Secondary* | 1.51 (0.93–2.45) | 1.19 (0.85–1.66) | 1.29 (0.98–1.69) | 2.38 (1.53–3.69) | 2.05 (1.54–2.72) | 2.15 (1.69–2.73) |
| *Tertiary* | 1.78 (0.85–3.73) | 1.75 (0.98–3.13) | 1.76 (1.12–2.79) | 4.09 (2.19–7.64) | 3.43 (2.07–5.68) | 3.69 (2.49–5.47) |
| **BMI** | | | | | | |
| *Underweight* | - - | - - | 2.82 (0.29–28.0) | 1.29 (0.09–19.1) | - - | 2.68 (0.25–29.3) |
| *Normal* | 2.37 (1.04–5.39) | 1.27 (0.79–2.03) | 1.48 (0.99–2.22) | 4.03 (1.92–8.48) | 2.51 (1.74–3.63) | 2.80 (2.01–3.91) |
| *Overweight* | 1.04 (0.64–1.70) | 1.25 (0.88–1.77) | 1.18 (0.89–1.57) | 2.09 (1.37–3.21) | 1.85 (1.37–2.49) | 1.93 (1.51–2.47) |
| *Obese* | 2.24 (1.06–4.71) | 0.87 (0.59–1.29) | 1.09 (0.78–1.54) | 3.81 (2.00–7.26) | 1.25 (0.90–1.76) | 1.67 (1.24–2.23) |

p-tau217, phosphorylated tau at threonine 217; CU, cognitively unimpaired; MCI, mild cognitive impairment; RR, risk ratio; CI, confidence interval; *APOE*, apolipoprotein E; BMI, body mass index. RRs are given using the low p-tau217 range as reference. Estimates were obtained from a quasi-Poisson regression model with a log link, adjusted for age, sex, education, *APOE* genotype, and BMI, incorporating interaction terms between p-tau217 range, cognitive status and characteristics at baseline. No significant differences between sexes were found when evaluated by rate of risk ratios tests. Inverse probability weighting was applied to reduce bias due to selective participation. Plasma p-tau217 data was missing from one participant. Two participants had cognitive impairment other than MCI.

Table S3. Estimated absolute risk of all-cause dementia by baseline cognitive status, plasma p-tau217 range and age groups.

| Clinical Diagnosis at baseline | Plasma p-tau217 at baseline | Age, 70–74 | | | Age, 75–79 | | | Age, 80–84 | | | Age, 85–89 | | | Age, 90+ | | |
|---|---|---|---|---|---|---|---|---|---|---|---|---|---|---|---|---|
| | | N | Risk (95% CI) | | N | Risk (95% CI) | | N | Risk (95% CI) | | N | Risk (95% CI) | | N | Risk (95% CI) | |
| CU | < 0.40 | 1038 | 3.3 | (2.3–4.8) | 472 | 4.4 | (2.7–6.9) | 163 | 7.9 | (4.5–13.3) | 32 | 15.0 | (5.9–33.1) | 11 | 30.6 | (9.6–64.5) |
| | 0.40–0.63 | 377 | 4.4 | (2.7–7.1) | 268 | 8.2 | (5.3–12.4) | 122 | 11.4 | (6.6–18.9) | 40 | 24.7 | (12.8–42.2) | 4 | - | - |
| | ≥ 0.63 | 220 | 8.9 | (5.7–13.7) | 216 | 11.8 | (8.0–17.1) | 141 | 24.1 | (17.5–32.3) | 52 | 35.2 | (22.6–50.3) | 14 | 55.1 | (26.7–80.6) |
| | All | 1635 | 4.3 | (3.4–5.5) | 956 | 7.1 | (5.6–9.1) | 426 | 14.4 | (11.2–18.3) | 124 | 26.1 | (18.7–35.2) | 29 | 37.1 | (20.9–56.7) |
| MCI | < 0.40 | 519 | 13.8 | (10.8–17.4) | 246 | 24.6 | (19.2–30.9) | 73 | 28.3 | (18.5–40.8) | 12 | 63.1 | (31.0–86.7) | 5 | 53.9 | (7.9–94.1) |
| | 0.40–0.63 | 237 | 15.6 | (11.3–21.2) | 149 | 25.1 | (18.3–33.6) | 68 | 40.0 | (28.3–52.9) | 28 | 66.0 | (45.3–81.9) | 5 | 69.7 | (14.7–96.8) |
| | ≥ 0.63 | 180 | 37.1 | (30.0–44.8) | 136 | 38.0 | (29.9–46.8) | 85 | 56.4 | (45.2–66.9) | 44 | 79.1 | (63.2–89.3) | 12 | 85.1 | (50.7–97.0) |
| | All | 936 | 18.9 | (16.4–21.8) | 531 | 28.3 | (24.4–32.6) | 226 | 42.8 | (36–49.8) | 84 | 72.3 | (61.2–81.2) | 22 | 72.9 | (48.9–88.4) |
| Total | < 0.40 | 1557 | 7.3 | (5.9–8.9) | 718 | 12.1 | (9.6–15.1) | 236 | 15.2 | (10.8–21.1) | 44 | 31.8 | (18.5–48.9) | 16 | 40.6 | (17.7–68.5) |
| | 0.40–0.63 | 614 | 9.1 | (6.9–11.8) | 417 | 14.9 | (11.5–19.1) | 190 | 23.2 | (17.2–30.5) | 68 | 45.6 | (33.2–58.6) | 9 | 45.3 | (13.7–81.3) |
| | ≥ 0.63 | 401 | 23.1 | (18.9–27.8) | 352 | 23.3 | (19.0–28.4) | 226 | 38.2 | (31.6–45.3) | 96 | 60.1 | (49.3–69.9) | 26 | 70.9 | (49.6–85.8) |
| | All | 2572 | 10.3 | (9.1–11.7) | 1488 | 15.6 | (13.7–17.7) | 652 | 25.9 | (22.3–29.7) | 208 | 49.2 | (42.0–56.5) | 51 | 55.9 | (41.0–69.7) |

p-tau217, phosphorylated tau at threonine 217; CI, confidence interval; CU, cognitively unimpaired; MCI, mild cognitive impairment; Concentration is given in picograms per milliliter. Age is given in years. Plasma p-tau217 data was missing from one participant. Two participants had cognitive impairment other than MCI. Estimates are weighted.

Table S4. Estimated absolute risk of all-cause dementia by baseline cognitive status, plasma p-tau217 range and *APOE* genotype.

| Clinical Diagnosis at baseline | Plasma p-tau217 at baseline | ε3ε3/ε3ε2/ε2ε2 | | | ε2ε4/ε3ε4 | | | ε4ε4 | | |
|---|---|---|---|---|---|---|---|---|---|---|
| | | N | Risk (95% CI) | | N | Risk (95% CI) | | N | Risk (95% CI) | |
| CU | < 0.40 | 1391 | 5.1 | (3.9–6.5) | 305 | 3.8 | (2.1–6.9) | 14 | - | - |
| | 0.40–0.63 | 528 | 8.7 | (6.3–11.9) | 266 | 7.0 | (4.4–11.0) | 14 | 6.9 | (0.8–41.1) |
| | ≥ 0.63 | 374 | 15.4 | (11.7–20) | 240 | 20.0 | (15.1–26.1) | 27 | 33.4 | (17.5–54.3) |
| | All | 2293 | 7.8 | (6.6–9.1) | 811 | 9.8 | (7.8–12.3) | 55 | 18.1 | (9.7–31.1) |
| MCI | < 0.40 | 694 | 21.1 | (17.8–24.7) | 154 | 17.0 | (11.2–25.1) | 2 | - | - |
| | 0.40–0.63 | 308 | 29.8 | (24.2–35.9) | 157 | 27.9 | (20.4–36.8) | 16 | 31.5 | (12.6–59.5) |
| | ≥ 0.63 | 224 | 50.6 | (43.5–57.6) | 192 | 47.0 | (39.6–54.6) | 36 | 56.3 | (38.5–72.7) |
| | All | 1226 | 29.6 | (26.8–32.6) | 503 | 32.3 | (27.9–37.0) | 54 | 47.6 | (33.7–62.0) |
| Total | < 0.40 | 2085 | 11.0 | (9.5–12.7) | 459 | 8.8 | (6.2–12.4) | 16 | - | - |
| | 0.40–0.63 | 836 | 17.4 | (14.5–20.7) | 423 | 15.9 | (12.1–20.6) | 30 | 20.5 | (9.1–39.9) |
| | ≥ 0.63 | 598 | 30.9 | (26.8–35.3) | 433 | 33.6 | (28.9–38.7) | 63 | 47.5 | (34.5–60.8) |
| | All | 3520 | 16.4 | (15.0–17.9) | 1315 | 19.7 | (17.3–22.3) | 109 | 33.7 | (24.8–44.0) |

p-tau217, phosphorylated tau at threonine 217; *APOE*, apolipoprotein E; CI, confidence interval; CU, cognitively unimpaired; MCI, mild cognitive impairment; Concentration is given in picograms per milliliter. Plasma p-tau217 data was missing from one participant. *APOE* genotype was missing from 11 CU and 16 MCI participants. Two participants had cognitive impairment other than MCI. Estimates are weighted.

Table S5. Estimated absolute risk of all-cause dementia by baseline cognitive status, plasma p-tau217 range and educational level.

| Clinical Diagnosis at baseline | Plasma p-tau217 at baseline | Primary education | | | Secondary education | | | Tertiary education | | |
|---|---|---|---|---|---|---|---|---|---|---|
| | | N | Risk (95% CI) | | N | Risk (95% CI) | | N | Risk (95% CI) | |
| CU | < 0.40 | 326 | 6.3 | (4.0–9.7) | 784 | 5.1 | (3.7–7.0) | 606 | 2.6 | (1.5–4.3) |
| | 0.40–0.63 | 169 | 9.1 | (5.4–15.0) | 373 | 9.0 | (6.3–12.7) | 268 | 5.4 | (3.2–9.0) |
| | ≥ 0.63 | 151 | 18.9 | (13.1–26.4) | 269 | 17.8 | (13.4–23.2) | 223 | 16.3 | (11.9–22.0) |
| | All | 646 | 10.1 | (7.9–12.9) | 1426 | 8.7 | (7.2–10.4) | 1097 | 6.2 | (4.8–7.9) |
| MCI | < 0.40 | 220 | 29.5 | (23.5–36.3) | 416 | 17.2 | (13.8–21.4) | 219 | 9.1 | (5.8–14.1) |
| | 0.40–0.63 | 115 | 39.1 | (30.1–49.0) | 239 | 25.4 | (20.0–31.8) | 131 | 16.5 | (10.8–24.4) |
| | ≥ 0.63 | 146 | 56.7 | (48.3–64.7) | 208 | 46.3 | (39.4–53.3) | 102 | 37.7 | (28.6–47.7) |
| | All | 481 | 40.7 | (36.2- 45.4) | 863 | 27.1 | (24.1–30.3) | 452 | 18.0 | (14.6- 22.0) |
| Total | < 0.40 | 546 | 16.6 | (13.4–20.3) | 1200 | 9.7 | (8.0–11.6) | 825 | 4.4 | (3.1–6.2) |
| | 0.40–0.63 | 284 | 23.0 | (18.0–29.0) | 612 | 15.9 | (13.0–19.4) | 399 | 9.3 | (6.7–12.8) |
| | ≥ 0.63 | 297 | 39.6 | (33.8–45.6) | 477 | 31.3 | (27.1–35.9) | 326 | 23.7 | (19.2–28.8) |
| | All | 1127 | 24.7 | (22.1–27.6) | 2290 | 16.2 | (14.7–17.9) | 1550 | 9.9 | (8.4–11.6) |

p-tau217, phosphorylated tau at threonine 217; CI, confidence interval; CU, cognitively unimpaired; MCI, mild cognitive impairment; Concentration is given in picograms per milliliter. Plasma p-tau217 data was missing from one participant. Education status was missing from four participants. Two participants had cognitive impairment other than MCI. Estimates are weighted. "Primary education" was defined ≤10 years of compulsory primary and lower secondary education; "Secondary education" was defined as additional 1–2 years of academic or vocational school, 3 years of academic or vocational school, or 3–4 years of vocational training or apprenticeship (upper secondary education); "Tertiary education" referred to college or university education.

Table S6. Estimated absolute risk of all-cause dementia by baseline cognitive status, plasma p-tau217 range and body mass index.

| Clinical Diagnosis at baseline | Plasma p-tau217 at baseline | BMI, Underweight | | | BMI, Normal | | | BMI, Overweight | | | BMI, Obese | | |
|---|---|---|---|---|---|---|---|---|---|---|---|---|---|
| | | N | Risk (95% CI) | | N | Risk (95% CI) | | N | Risk (95% CI) | | N | Risk (95% CI) | |
| CU | < 0.40 | 7 | 14.2 | (1.2–70.2) | 476 | 2.7 | (1.4–5.1) | 826 | 5.4 | (4.0–7.4) | 392 | 4.0 | (2.3–6.8) |
| | 0.40–0.63 | 2 | - | - | 245 | 7.6 | (4.8–12.0) | 400 | 7.4 | (5.0–10.9) | 160 | 9.9 | (5.7–16.6) |
| | ≥ 0.63 | 5 | 15.3 | (0.8–80.3) | 216 | 15.1 | (10.6–21.1) | 286 | 17.9 | (13.4–23.4) | 126 | 20.3 | (13.7–29.1) |
| | All | 14 | 12.4 | (2.6–42.9) | 937 | 7.0 | (5.4–9.1) | 1512 | 8.5 | (7.1–10.2) | 678 | 8.6 | (6.5–11.3) |
| MCI | < 0.40 | 2 | - | - | 202 | 17.7 | (12.4–24.7) | 410 | 17.7 | (13.7–22.5) | 232 | 25.7 | (20.1–32.2) |
| | 0.40–0.63 | 6 | 33.1 | (4.6–83.5) | 135 | 23.4 | (16.4–32.2) | 208 | 26.0 | (19.7–33.6) | 114 | 27.6 | (19.3–38.0) |
| | ≥ 0.63 | 5 | 15.3 | (0.8–80.6) | 154 | 55.5 | (47.1–63.6) | 183 | 43.4 | (35.8–51.3) | 93 | 43.7 | (33.1–54.8) |
| | All | 13 | 21.6 | (5.7–55.6) | 491 | 32.4 | (27.9–37.2) | 801 | 26.3 | (23.0–30.0) | 439 | 30.4 | (25.9–35.4) |
| Total | < 0.40 | 9 | 10.8 | (1.0–58.5) | 678 | 7.8 | (5.6–10.6) | 1236 | 10.1 | (8.2–12.3) | 624 | 12.7 | (10.0–15.9) |
| | 0.40–0.63 | 8 | 25.3 | (4.3–72.0) | 380 | 13.8 | (10.4–18.2) | 608 | 14.5 | (11.5–18.2) | 274 | 18.0 | (13.3–24.0) |
| | ≥ 0.63 | 10 | 15.3 | (2.9–52.3) | 370 | 34.4 | (29.2–40.0) | 470 | 29.4 | (25.0–34.3) | 219 | 31.4 | (25.0–38.6) |
| | All | 27 | 17.1 | (6.6–37.5) | 1428 | 17.0 | (14.8–19.4) | 2315 | 15.6 | (13.9–17.4) | 1117 | 18.0 | (15.6–20.7) |

p-tau217, phosphorylated tau at threonine 217; BMI, body mass index; CI, confidence interval; CU, cognitively unimpaired; MCI, mild cognitive impairment; Concentration is given in picograms per milliliter. Plasma p-tau217 data was missing from one participant. BMI data was missing from 29 CU participants and 55 MCI participants. Two participants had cognitive impairment other than MCI. Estimates are weighted.

#### Supplementary table on odds associated with continuous biomarker measurements

Table S7. Odds Ratios for all-cause dementia associated with continuous plasma p-tau217 and NfL and clinical covariates from a Survey-Weighted Logistic Regression Model.

| Variable | OR | 95% CI | *P* value |
|---|---|---|---|
| **Plasma p-tau217 (per pg/mL)** | 4.53 | 2.94–6.98 | <.0001 |
| **Cognitive group** | | | |
| *MCI vs CU* | 6.56 | 4.52–9.52 | <.0001 |
| **Sex** | | | |
| *Male vs Female* | 1.91 | 1.31–2.80 | .0008 |
| **Education** | | | |
| *Secondary vs Primary* | 1.07 | 0.74–1.53 | .709 |
| *Tertiary vs Primary* | 0.79 | 0.52–1.23 | .251 |
| **Plasma NfL** | Natural spline (2 df) | — | .0100 |
| **Serum creatinine** | 1 | 0.99–1.00 | .465 |
| **APOE ε4** | | | |
| *1 allele vs 0* | 1.27 | 1.02–1.58 | .031 |
| *2 alleles vs 0* | 2.62 | 1.58–4.37 | .0002 |
| **Age group** | | | |
| *75–80 vs <75* | 1.52 | 1.20–1.93 | <.0001 |
| *80–85 vs <75* | 2.48 | 1.85–3.33 | <.0001 |
| *85–90 vs <75* | 6.29 | 4.20–9.42 | <.0001 |
| *≥90 vs <75* | 5.56 | 2.51–12.34 | <.0001 |
| **BMI** | | | |
| *Normal vs Underweight* | 0.96 | 0.33–2.80 | .946 |
| *Overweight vs Underweight* | 0.92 | 0.32–2.72 | .882 |
| *Obese vs Underweight* | 1.31 | 0.45–3.87 | .62 |
| **Interactions** | | | |
| *Plasma p-tau217 × Male* | 0.34 | 0.19–0.61 | <.0001 |
| *MCI × Secondary education* | 0.6 | 0.38–0.95 | .032 |
| *MCI × Tertiary education* | 0.55 | 0.33–0.93 | .027 |

p-tau217, phosphorylated tau at threonine 217; NfL, neurofilament light; OR, odds ratio; CI, confidence interval; CU, cognitively unimpaired; MCI, mild cognitive impairment; *APOE*, apolipoprotein E; BMI, Body mass index. Associations were estimated using survey-weighted logistic regression. Given the weighted cumulative incidence of dementia (17.7%), ORs may moderately overestimate prevalence ratios. A survey-weighted log-binomial model was explored but exhibited numerical instability; therefore, the logistic regression model was retained as the primary analysis.

#### Supplementary tables on NfL and all-cause dementia

Table S8. Estimated absolute risk of all-cause dementia by baseline cognitive status, plasma NfL range and sex.

| Clinical Diagnosis at baseline | Plasma NfL at baseline | Female | | | Male | | | Total | | |
|---|---|---|---|---|---|---|---|---|---|---|
| | | N | Risk (95% CI) | | N | Risk (95% CI) | | N | Risk (95% CI) | |
| CU | < cut-off | 1583 | 8.0 | (6.6–9.6) | 1295 | 6.5 | (5.2–8.2) | 2878 | 7.3 | (6.3–8.5) |
| | ≥ cut-off | 140 | 20.3 | (13.8 -28.9) | 119 | 21.0 | (14.0–30.2) | 259 | 20.6 | (15.6–26.7) |
| | All | 1746 | 9.0 | (7.6–10.7) | 1424 | 7.8 | (6.4–9.5) | 3170 | 8.5 | (7.5–9.6) |
| MCI | < cut-off | 779 | 30.8 | (27.2–34.6) | 792 | 25.4 | (22.2–28.9) | 1571 | 28.2 | (25.7–30.8) |
| | ≥ cut-off | 103 | 53.6 | (43.1–63.7) | 95 | 45.9 | (35.3–56.9) | 198 | 50.3 | (42.8–57.8) |
| | All | 894 | 33.8 | (30.4–37.5) | 905 | 27.6 | (24.5–31.0) | 1799 | 30.9 | (28.5–33.3) |
| Total | < cut-off | 2364 | 16.6 | (14.9–18.5) | 2087 | 14.5 | (12.9–16.3) | 4451 | 15.7 | (14.4–17.0) |
| | ≥ cut-off | 243 | 37.0 | (30.4–41.2) | 214 | 33.3 | (26.6–40.7) | 457 | 35.4 | (30.6–40.5) |
| | All | 2642 | 18.8 | (17.1–20.7) | 2329 | 16.4 | (14.7–18.2) | 4971 | 17.7 | (16.5–19.0) |

NfL, neurofilament light; CI, confidence interval; CU, cognitively unimpaired; MCI, mild cognitive impairment; Plasma NfL data were missing from 35 females and 28 males. Two participants had cognitive impairment other than MCI. Estimates are weighted.

Table S9. Association between elevated plasma neurofilament light range at baseline and risk of all-cause dementia at follow-up across demographic and clinical subgroups (N, 4,795).

| **Characteristics at baseline** | **CU** | | **MCI** | | **Total** | |
|---|---|---|---|---|---|---|
| | **RR (95% CI)** | | **RR (95% CI)** | | **RR (95% CI)** | |
| All | 1.78 | (1.28–2.49) | 1.70 | (1.15–2.51) | 1.72 | (1.31–2.27) |
| Sex | | | | | | |
| *Female* | 1.78 | (1.179–2.69) | 1.80 | (0.984–3.29) | 1.79 | (1.17–2.74) |
| *Male* | 1.79 | (1.074–2.97) | 1.58 | (1.197–2.08) | 1.63 | (1.28–2.08) |
| Age | | | | | | |
| *70–74* | 2.01 | (0.878–4.58) | 2.70 | (1.532–4.76) | 2.52 | (1.615–3.92) |
| *75–79* | 1.29 | (0.505–3.3) | 1.42 | (0.725–2.76) | 1.38 | (0.795–2.4) |
| *80–84* | 2.01 | (1.158–3.48) | 1.41 | (0.909–2.18) | 1.62 | (1.151–2.28) |
| *85–89* | 1.75 | (0.916–3.33) | 1.16 | (0.809–1.65) | 1.31 | (0.96–1.79) |
| *90+* | 2.50 | (0.532–11.72) | 0.63 | (0.242–1.63) | 0.99 | (0.491–2) |
| *APOE* genotype | | | | | | |
| *ε3ε3/ε3ε2/ε2ε2* | 1.59 | (1.046–2.41) | 1.49 | (1.024–2.17) | 1.52 | (1.16–2) |
| *ε2ε4/ε3ε4* | 2.13 | (1.248–3.64) | 1.95 | (1.14–3.32) | 2.00 | (1.33–2.98) |
| *ε4ε4* | 2.61 | (0.933–7.31) | 3.05 | (1.523–6.1) | 2.93 | (1.64–5.25) |
| Education | | | | | | |
| *Primary* | 1.75 | (0.968–3.17) | 1.43 | (0.947–2.16) | 1.50 | (1.08–2.09) |
| *Secondary* | 1.54 | (0.95–2.49) | 1.80 | (1.135–2.86) | 1.72 | (1.23–2.39) |
| *Tertiary* | 2.46 | (1.447–4.18) | 2.71 | (1.512–4.86) | 2.60 | (1.72–3.94) |
| BMI | | | | | | |
| *Underweight* | - | - | - | - | - | - |
| *Normal* | 1.55 | (0.983–2.45) | 1.35 | (0.998–1.84) | 1.4 | (1.04–1.89) |
| *Overweight* | 1.75 | (1.177–2.59) | 1.5 | (1.141–1.98) | 1.58 | (1.23–2.03) |
| *Obese* | 1.67 | (1.031–2.72) | 1.59 | (1.073–2.35) | 1.61 | (1.12–2.32) |

CU, cognitively unimpaired; MCI, mild cognitive impairment; RR, risk ratio; CI, confidence interval; *APOE*, apolipoprotein E; BMI, body mass index. RRs are given using the low neurofilament light (NfL) range as reference. Estimates were obtained from a quasi-Poisson regression model with a log link, adjusted for age, sex, education, *APOE* genotype, and BMI, incorporating interaction terms between NfL range, cognitive status and characteristics at baseline. No significant differences between sexes were found when evaluated by rate of risk ratios tests. Inverse probability weighting was applied to reduce bias due to selective participation. Plasma NfL data were missing from 63 participants. Two participants had cognitive impairment other than MCI.

Table S10. Estimated absolute risk of all-cause dementia by baseline cognitive status, plasma NfL range and age groups.

| Clinical Diagnosis at baseline | Plasma NfL at baseline | Age, 70–74 | | | Age, 75–79 | | | Age, 80–84 | | | Age, 85–89 | | | Age, 90+ | | |
|---|---|---|---|---|---|---|---|---|---|---|---|---|---|---|---|---|
| | | N | Risk (95%CI) | | N | Risk (95%CI) | | N | Risk (95%CI) | | N | Risk (95%CI) | | N | Risk (95%CI) | |
| CU | < cut-off | 1501 | 4.0 | (3.1–5.2) | 897 | 6.8 | (5.3–8.8) | 373 | 12.9 | (9.7–17.0) | 93 | 22.6 | (14.8–33.0) | 14 | 25.7 | (8.6–56.0) |
| | ≥ cut-off | 111 | 8.0 | (3.8–16.3) | 53 | 12.8 | (5.7–26.5) | 50 | 26.1 | (15.5–40.6) | 31 | 36.1 | (20.1–55.9) | 14 | 54.6 | (26.4–80.1) |
| | All | 1635 | 4.3 | (3.4–5.5) | 956 | 7.1 | (5.6–9.1) | 426 | 14.4 | (11.2–18.3) | 124 | 26.1 | (18.7–35.2) | 29 | 37.1 | (20.9–56.7) |
| MCI | < cut-off | 841 | 16.8 | (14.3–19.7) | 483 | 28.2 | (24.1–32.7) | 181 | 40.5 | (33.1–48.4) | 56 | 74.1 | (60.3–84.3) | 10 | 82 | (41.6–96.7) |
| | ≥ cut-off | 78 | 40.1 | (29.2–52.0) | 39 | 31.6 | (18.1–49.3) | 43 | 51.8 | (36.1–67.2) | 26 | 73.9 | (51.3–88.4) | 12 | 64.9 | (31.3–88.2) |
| | All | 936 | 18.9 | (16.4–21.8) | 531 | 28.3 | (24.4–32.6) | 226 | 42.8 | (36.0–49.8) | 84 | 72.3 | (61.2–81.2) | 22 | 72.9 | (48.9–88.4) |
| Total | < cut-off | 2343 | 9.2 | (8.0–10.5) | 1381 | 15.2 | (13.2–17.4) | 554 | 23.5 | (19.8–27.7) | 149 | 47.1 | (38.6–55.8) | 24 | 54.1 | (32.6–74.1) |
| | ≥ cut-off | 189 | 22.7 | (16.8–29.9) | 92 | 21.9 | (13.9–32.7) | 93 | 39.4 | (29.3–50.6) | 57 | 56.5 | (42.4–69.6) | 26 | 60.5 | (38.8–78.7) |
| | All | 2572 | 10.3 | (9.1–11.7) | 1488 | 15.6 | (13.7–17.7) | 652 | 25.9 | (22.3–29.7) | 208 | 49.2 | (42.0–56.5) | 51 | 55.9 | (41.0–69.7) |

NfL, neurofilament light; CI, confidence interval; CU, cognitively unimpaired; MCI, mild cognitive impairment. Age is given in years. Plasma NfL data were missing from 63 participants. Two participants had cognitive impairment other than MCI. Estimates are weighted.

Table S11. Estimated absolute risk of all-cause dementia by baseline cognitive status, plasma NfL range and *APOE* genotype.

| Clinical Diagnosis at baseline | Plasma NfL at baseline | ε3ε3/ε3ε2/ε2ε2 | | | ε2ε4/ε3ε4 | | | ε4ε4 | | |
|---|---|---|---|---|---|---|---|---|---|---|
| | | N | Risk (95% CI) | | N | Risk (95% CI) | | N | Risk (95% CI) | |
| CU | < cut-off | 2084 | 7.0 | (5.8–8.3) | 733 | 7.7 | (5.9–10.0) | 50 | 15.3 | (7.6–28.5) |
| | ≥ cut-off | 185 | 15.9 | (10.9–22.7) | 69 | 31.4 | (20.5–44.9) | 5 | 49.6 | (7.1–92.7) |
| | All | 2293 | 7.8 | (6.6–9.1) | 811 | 9.8 | (7.8–12.3) | 55 | 18.1 | (9.7–31.1) |
| MCI | < cut-off | 1067 | 26.8 | (23.8–29.9) | 442 | 30.1 | (25.5–35.1) | 47 | 40.9 | (26.7–56.9) |
| | ≥ cut-off | 135 | 49.5 | (40.4–58.6) | 56 | 50.0 | (35.9–64.1) | 6 | 87.7 | (29.3–99.2) |
| | All | 1226 | 29.6 | (26.8–32.6) | 503 | 32.3 | (27.9–37.0) | 54 | 47.6 | (33.7–62.0) |
| Total | < cut-off | 3152 | 14.5 | (13.1–16) | 1176 | 17.4 | (15.0–20.0) | 97 | 28.5 | (19.7–39.3) |
| | ≥ cut-off | 320 | 32.6 | (27.0–38.8) | 125 | 40.5 | (31.4–50.4) | 11 | 71.8 | (38.0–91.4) |
| | All | 3520 | 16.4 | (15.0–17.9) | 1315 | 19.7 | (17.3–22.3) | 109 | 33.7 | (24.8–44.0) |

NfL, neurofilament light; *APOE*, apolipoprotein E; CI, confidence interval; CU, cognitively unimpaired; MCI, mild cognitive impairment; Plasma NfL data were missing from 63 participants. *APOE* genotype was missing for 11 CU and 16 MCI participants. Two participants had cognitive impairment other than MCI. Estimates are weighted.

Table S12. Estimated absolute risk of all-cause dementia by baseline cognitive status, plasma NfL range and educational level.

| Clinical Diagnosis at baseline | Plasma NfL at baseline | Primary education | | | Secondary education | | | Tertiary education | | |
|---|---|---|---|---|---|---|---|---|---|---|
| | | N | Risk (95% CI) | | N | Risk (95% CI) | | N | Risk (95% CI) | |
| CU | < cut-off | 577 | 8.8 | (6.7–11.6) | 1300 | 7.7 | (6.3–9.4) | 1001 | 5.0 | (3.8–6.6) |
| | ≥ cut-off | 59 | 22.8 | (13.3–36.3) | 108 | 18.8 | (12.1–28.1) | 91 | 19.1 | (11.8–29.3) |
| | All | 646 | 10.1 | (7.9–12.9) | 1426 | 8.7 | (7.2–10.4) | 1097 | 6.2 | (4.8–7.9) |
| MCI | < cut-off | 417 | 38.2 | (33.4–43.2) | 747 | 24.6 | (21.5–28.0) | 406 | 15.3 | (12.0–19.4) |
| | ≥ cut-off | 57 | 58.6 | (45.0–71.0) | 98 | 44.1 | (34.2–54.5) | 41 | 44.3 | (29.4–60.3) |
| | All | 481 | 40.7 | (36.2–45.4) | 863 | 27.1 | (24.1–30.3) | 452 | 18.0 | (14.6–22.0) |
| Total | < cut-off | 994 | 22.5 | (19.8–25.5) | 2048 | 14.4 | (12.9–16.1) | 1408 | 8.2 | (6.8–9.9) |
| | ≥ cut-off | 116 | 42.6 | (33.4–52.4) | 206 | 31.9 | (25.5–39.0) | 132 | 27.5 | (20.2–36.3) |
| | All | 1127 | 24.7 | (22.1–27.6) | 2290 | 16.2 | (14.7–17.9) | 1550 | 9.9 | (8.4–11.6) |

NfL, neurofilament light; CI, confidence interval; CU, cognitively unimpaired; MCI, mild cognitive impairment; Plasma NfL data were missing from 63 participants. Education status was missing for four participants. Two participants had cognitive impairment other than MCI. Estimates are weighted. "Primary education" was defined ≤10 years of compulsory primary and lower secondary education; "Secondary education" was defined as additional 1–2 years of academic or vocational school, 3 years of academic or vocational school, or 3–4 years of vocational training or apprenticeship (upper secondary education); "Tertiary education" referred to college or university education.

Table S13. Estimated absolute risk of all-cause dementia by baseline cognitive status, plasma NfL range and body mass index.

| Clinical Diagnosis at baseline | Plasma NfL at baseline | BMI, Underweight | | | BMI, Normal | | | BMI, Overweight | | | BMI, Obese | | |
|---|---|---|---|---|---|---|---|---|---|---|---|---|---|
| | | N | Risk (95% CI) | | N | Risk (95% CI) | | N | Risk (95% CI) | | N | Risk (95% CI) | |
| CU | < cut-off | 13 | 5.7 | (0.6–36.8) | 840 | 6.0 | (4.5–8.1) | 1374 | 7.6 | (6.2–9.3) | 629 | 7.4 | (5.5–10.0) |
| | ≥ cut-off | 1 | - | - | 90 | 16.3 | (9.5–26.6) | 117 | 18.4 | (11.7–27.9) | 44 | 23.8 | (12.3–41.0) |
| | All | 14 | 12.4 | (2.6–42.9) | 937 | 7 | (5.4–9.1) | 1512 | 8.5 | (7.1–10.2) | 678 | 8.6 | (6.5–11.3) |
| MCI | < cut-off | 13 | 21.6 | (5.7–55.6) | 420 | 30.4 | (25.7–35.6) | 698 | 23.5 | (20.1–27.3) | 409 | 29.4 | (24.7–34.5) |
| | ≥ cut-off | - | - | - | 62 | 44.3 | (31.3–58.1) | 87 | 47.3 | (36.1–58.8) | 26 | 47.3 | (27.7–67.7) |
| | All | 13 | 21.6 | (5.7–55.6) | 491 | 32.4 | (27.9–37.2) | 801 | 26.3 | (23–30) | 439 | 30.4 | (25.9–35.4) |
| Total | < cut-off | 26 | 14.1 | (4.8–34.9) | 1260 | 15.3 | (13.1–17.8) | 2074 | 13.7 | (12.1–15.6) | 1038 | 16.9 | (14.5–19.7) |
| | ≥ cut-off | 1 | - | - | 152 | 29.2 | (21.7–38) | 204 | 32.8 | (25.9–40.6) | 70 | 32.7 | (21.8–45.7) |
| | All | 27 | 17.1 | (6.6–37.5) | 1428 | 17.0 | (14.8–19.4) | 2315 | 15.6 | (13.9–17.4) | 1117 | 18.0 | (15.6–20.7) |

NfL, neurofilament light; BMI, Body mass index; CI, confidence interval; CU, cognitively unimpaired; MCI, mild cognitive impairment; Plasma NfL data were missing from 63 participants. BMI data were missing from 29 CU participants and 55 MCI participants. Two participants had cognitive impairment other than MCI. Estimates are weighted.

#### Supplementary tables on joint p-tau217 and NfL and all-cause dementia

Table S14. Estimated absolute risk of all-cause dementia by baseline cognitive status, plasma p-tau217 and NfL range and sex.

| Clinical Diagnosis at baseline | Plasma p-tau217 and NfL at baseline | Female | | | Male | | | Total | | |
|---|---|---|---|---|---|---|---|---|---|---|
| | | N | Risk | (95% CI) | N | Risk | (95% CI) | N | Risk | (95% CI) |
| CU | < cut-off | 935 | 4.6 | (3.3–6.4) | 680 | 4.5 | (3.1–6.5) | 1615 | 4.6 | (3.6–5.9) |
| | ≥ cut-off | 55 | 26.7 | (16.0–41.1) | 61 | 32.2 | (20.8–46.1) | 116 | 29.4 | (21.3–39.1) |
| | All | 1746 | 9.0 | (7.6–10.7) | 1424 | 7.8 | (6.4–9.5) | 3170 | 8.5 | (7.5–9.6) |
| MCI | < cut-off | 398 | 22.3 | (17.9–27.4) | 392 | 18.6 | (14.8–23.2) | 790 | 20.5 | (17.5–23.9) |
| | ≥ cut-off | 52 | 66.7 | (51.9–78.8) | 49 | 61.0 | (45.6–74.5) | 101 | 64.2 | (53.8–73.5) |
| | All | 894 | 33.8 | (30.4–37.5) | 905 | 27.6 | (24.5–31.0) | 1799 | 30.9 | (28.5–33.3) |
| Total | < cut-off | 1333 | 10.6 | (8.8–12.8) | 1072 | 10.3 | (8.4–12.5) | 2405 | 10.5 | (9.1–12.0) |
| | ≥ cut-off | 107 | 48.9 | (38.8–59.1) | 110 | 46.5 | (36.6–56.7) | 217 | 47.8 | (40.7–55.0) |
| | All | 2642 | 19 | (17.1–20.7) | 1147 | 16.4 | (14.7–18.2) | 3789 | 17.7 | (16.5–19.0) |

p-tau217, phosphorylated tau at threonine 217; NfL, neurofilament light; CI, confidence interval; CU, cognitively unimpaired; MCI, mild cognitive impairment; Plasma p-tau217 data was missing from one participant. Plasma NfL data were missing from 35 females and 28 males. Two participants had cognitive impairment other than MCI. Estimates are weighted.

Table 15. Association between baseline joint high versus low p-tau217 and NfL range and risk of all-cause dementia at follow-up across demographic and clinical subgroups (N, 2,572).

| **Characteristics at baseline** | **CU** | | **MCI** | | **Total** | |
|---|---|---|---|---|---|---|
| | **RR (95% CI)** | | **RR (95% CI)** | | **RR (95% CI)** | |
| All | 3.07 | (1.64–5.74) | 3.41 | (2.65–4.38) | 3.32 | (2.61–4.23) |
| Sex | | | | | | |
| *Female* | 3.33 | (1.63–6.82) | 3.10 | (2.26–4.25) | 3.16 | (2.34–4.25) |
| *Male* | 2.75 | (1.21–6.22) | 3.78 | (2.61–5.47) | 3.52 | (2.52–4.93) |
| Age | | | | | | |
| *70–74* | 3.33 | (0.845–13.08) | 7.37 | (5.265–10.32) | 6.28 | (4.457–8.84) |
| *75–79* | 2.06 | (0.65–6.53) | 1.77 | (1.011–3.1) | 1.83 | (1.103–3.05) |
| *80–84* | 3.82 | (1.691–8.64) | 1.67 | (0.902–3.08) | 2.36 | (1.419–3.91) |
| *85–89* | 2.58 | (0.758–8.81) | 1.10 | (0.648–1.87) | 1.40 | (0.859–2.27) |
| *90+* | 5.33 | (0.644–44.1) | 1.34 | (0.53–3.37) | 1.93 | (0.873–4.27) |
| *APOE* genotype | | | | | | |
| *ε3ε3/ε3ε2/ε2ε2* | 2.20 | (1.08–4.5) | 3.28 | (2.4–4.48) | 3.02 | (2.27–4.02) |
| *ε2ε4/ε3ε4* | 6.08 | (2.44–15.2) | 3.85 | (2.47–6.01) | 4.46 | (2.88–6.9) |
| *ε4ε4* | - | - | - | - | - | - |
| Education | | | | | | |
| *Primary* | 2.37 | (1.04–5.39) | 2.71 | (1.86–3.94) | 2.65 | (1.88–3.73) |
| *Secondary* | 2.99 | (1.33–6.7) | 3.37 | (2.41–4.73) | 3.26 | (2.35–4.53) |
| *Tertiary* | 5.07 | (2.09–12.29) | 8.50 | (4.82–15) | 7.17 | (4.48–11.47) |
| BMI | | | | | | |
| *Underweight* | - | - | - | - | - | - |
| *Normal* | 3.27 | (1.788–5.99) | 3.34 | (2.579–4.34) | 3.33 | (2.616–4.24) |
| *Overweight* | 3.08 | (1.603–5.91) | 3.05 | (2.358–3.96) | 3.06 | (2.357–3.98) |
| *Obese* | 2.89 | (1.534–5.43) | 4.01 | (3.104–5.18) | 3.76 | (2.96–4.78) |

p-tau217, phosphorylated tau at threonine 217; NfL, neurofilament light; CU, cognitively unimpaired; MCI, mild cognitive impairment; RR, risk ratio; CI, confidence interval; *APOE*, apolipoprotein E; BMI, body mass index. RRs are given using the low p-tau217 and NfL ranges as reference. Estimates were obtained from a quasi-Poisson regression model with a log link, adjusted for age, sex, education, *APOE* genotype, and BMI, incorporating interaction terms between p-tau217 and NfL ranges, cognitive status and characteristics at baseline. No significant differences between sexes were found when evaluated by rate of risk ratios tests. Inverse probability weighting was applied to reduce bias due to selective participation.

Plasma p-tau217 data was missing from one participant. Plasma NfL data were missing from 63 participants. Two participants had cognitive impairment other than MCI.

Table S16. Estimated absolute risk of all-cause dementia by baseline cognitive status, joint plasma p-tau217 and NfL ranges and age groups.

| Clinical Diagnosis at baseline | Plasma p-tau217 and NfL at baseline | Age, 70–74 | | | Age, 75–79 | | | Age, 80–84 | | | Age, 85–89 | | | Age, 90+ | | |
|---|---|---|---|---|---|---|---|---|---|---|---|---|---|---|---|---|
| | | N | Risk | (95% CI) | N | Risk | (95% CI) | N | Risk | (95% CI) | N | Risk | (95% CI) | N | Risk | (95% CI) |
| CU | < cut-off | 966 | 3.1 | (2.1–4.5) | 457 | 4.5 | (2.8–7.1) | 154 | 7.7 | (4.3–13.2) | 29 | 14.7 | (5.3–34.6) | 9 | 29.0 | (7.3–68.2) |
| | ≥ cut-off | 27 | 10.8 | (2.5–37.0) | 24 | 11.9 | (3.6–32.9) | 32 | 33.4 | (18.8–52.0) | 23 | 35.3 | (17.4–58.4) | 10 | 70.0 | (31.2–92.3) |
| | All | 1635 | 4.3 | (3.4–5.5) | 956 | 7.1 | (5.6–9.1) | 426 | 14.4 | (11.2–18.3) | 124 | 26.1 | (18.7–35.2) | 29 | 37.1 | (20.9–56.7) |
| MCI | < cut-off | 476 | 13.9 | (10.8–17.7) | 235 | 25.3 | (19.7–31.8) | 67 | 28.5 | (18.3–41.5) | 8 | 71.0 | (29.1–93.6) | 4 | 72.3 | (5.8–99.1) |
| | ≥ cut-off | 21 | 92.1 | (70.9–98.2) | 24 | 36.9 | (18.8–59.6) | 29 | 54.6 | (35.3–72.7) | 18 | 73.7 | (45.0–90.6) | 9 | 79.9 | (38.2–96.3) |
| | All | 963 | 18.9 | (16.4–21.8) | 531 | 28.3 | (24.4–32.6) | 226 | 42.8 | (36.0–49.8) | 84 | 72.3 | (61.2–81.2) | 22 | 72.9 | (48.9–88.4) |
| Total | < cut-off | 1442 | 7.1 | (5.7–8.8) | 692 | 12.4 | (9.9–15.4) | 221 | 15.0 | (10.5–20.9) | 37 | 30.2 | (16.2–49.2) | 13 | 46.6 | (19.3–76.1) |
| | ≥ cut-off | 48 | 50.8 | (36.1–65.4) | 48 | 25.9 | (14.6–41.7) | 61 | 44.4 | (31.6–57.9) | 41 | 55.7 | (38.9–71.3) | 19 | 75.5 | (49.8 -90.5) |
| | All | 2572 | 10.3 | (9.1–11.7) | 1488 | 15.6 | (13.7–17.7) | 652 | 25.9 | (22.3–29.7) | 208 | 49.2 | (42.0–56.5) | 51 | 55.9 | (41.0–69.7) |

p-tau217, phosphorylated tau at threonine 217; NfL, neurofilament light; CI, confidence interval; CU, cognitively unimpaired; MCI, mild cognitive impairment; Age is given in years. Plasma p-tau217 data was missing from one participant. Plasma NfL data were missing from 63 participants. Two participants had cognitive impairment other than MCI. Estimates are weighted.

Table S17. Estimated absolute risk of all-cause dementia by baseline cognitive status, joint plasma p-tau217 and NfL range and *APOE* genotype.

| Clinical Diagnosis at baseline | Plasma p-tau217 and NfL at baseline | ε3ε3/ε3ε2/ε2ε2 | | | ε2ε4/ε3ε4 | | | ε4ε4 | | |
|---|---|---|---|---|---|---|---|---|---|---|
| | | N | Risk | (95% CI) | N | Risk | (95% CI) | N | Risk | (95% CI) |
| CU | < cut-off | 1309 | 4.8 | (3.6–6.3) | 286 | 4.1 | (2.2–7.3) | 14 | - | - |
| | ≥ cut-off | 72 | 17.7 | (10.2–28.8) | 40 | 48.5 | (32.3–65.0) | 4 | 58.6 | (5.4–97.2) |
| | All | 2293 | 7.8 | (6.6–9.1) | 811 | 9.8 | (7.8–12.3) | 55 | 18.1 | (9.7–31.1) |
| MCI | < cut-off | 641 | 21.3 | (17.9–25.1) | 142 | 17.3 | (11.1–25.8) | 2 | - | - |
| | ≥ cut-off | 61 | 65.1 | 51.5–76.6) | 35 | 59.9 | (41.7–75.8) | 5 | 85.1 | (19.6–99.3) |
| | All | 1226 | 29.6 | (26.8–32.6) | 503 | 32.3 | (27.9–37.0 | 54 | 47.6 | (33.7–62.0) |
| Total | < cut-off | 1950 | 10.8 | (9.3–12.6) | 428 | 9.1 | (6.3–12.9) | 16 | - | - |
| | ≥ cut-off | 133 | 43.1 | (34.2–52.6) | 75 | 54.3 | (42.1–65.9) | 9 | 73.9 | (34.9–93.7) |
| | All | 3520 | 16.4 | (15.0–17.9) | 1315 | 19.7 | (17.3–22.3) | 109 | 33.7 | (24.8–44.0) |

p-tau217, phosphorylated tau at threonine 217; NfL, neurofilament light; *APOE*, apolipoprotein E; CI, confidence interval; CU, cognitively unimpaired; MCI, mild cognitive impairment; Two participants had cognitive impairment other than MCI. Plasma p-tau217 data was missing from one participant. Plasma NfL data were missing from 63 participants. *APOE* genotype data was missing from 11 CU and 16 MCI participants. Estimates are weighted.

Table S18. Estimated absolute risk of all-cause dementia by baseline cognitive status, joint plasma p-tau217 and NfL range and educational level.

| Clinical Diagnosis at baseline | Plasma p-tau217 and NfL at baseline | Primary education | | | Secondary education | | | Tertiary education | | |
|---|---|---|---|---|---|---|---|---|---|---|
| | | **N** | **Risk (95% CI)** | | **N** | **Risk (95% CI)** | | **N** | **Risk (95% CI)** | |
| CU | < cut-off | 307 | 6.3 | (4.0–9.8) | 732 | 4.9 | (3.5–6.8) | 576 | 2.3 | (1.3–4.0) |
| | ≥ cut-off | 31 | 24.2 | (11. 7–43.6) | 42 | 34.2 | (20.8–50.8) | 43 | 30.8 | (18.3–47.0) |
| | All | 646 | 10.1 | (7.9–12.9) | 1426 | 8.7 | (7.2–10.4) | 478 | 6.2 | (4.8–7.9) |
| MCI | < cut-off | 204 | 30.0 | (23.7–37.0) | 382 | 17.6 | (14.0–21.9) | 204 | 8.8 | (5.4–13.9) |
| | ≥ cut-off | 29 | 73.6 | (53.6–87.0) | 49 | 57.4 | (42.7–70.8) | 22 | 64.7 | (42.0–82.3) |
| | All | 481 | 40.7 | (36.2–45.4) | 863 | 27.1 | (24.1–30.3) | 452 | 18.0 | (16.4–22.0) |
| Total | < cut-off | 511 | 16.6 | (13.4–20.4) | 1114 | 9.6 | (7.9–11.6) | 780 | 4.1 | (2.8–5.9) |
| | ≥ cut-off | 60 | 51.0 | (38.0–63.9) | 91 | 47.7 | (37.3–58.3) | 65 | 43.4 | (31.5–56.1) |
| | All | 1127 | 24.7 | (22.1–27.6) | 2290 | 16.2 | (14.7–17.9) | 1550 | 9.9 | (8.4–11.6) |

p-tau217, phosphorylated tau at threonine 217; NfL, neurofilament light; CI, confidence interval; CU, cognitively unimpaired; MCI, mild cognitive impairment; Two participants had cognitive impairment other than MCI. Plasma p-tau217 data was missing from one participant. Plasma NfL data were missing from 63 participants. Education status was missing from four participants. Estimates are weighted. "Primary education" was defined ≤10 years of compulsory primary and lower secondary education; "Secondary education" was defined as additional 1–2 years of academic or vocational school, 3 years of academic or vocational school, or 3–4 years of vocational training or apprenticeship (upper secondary education); "Tertiary education" referred to college or university education.

Table S19. Estimated absolute risk of all-cause dementia by baseline cognitive status, joint plasma p-tau217 and NfL range and body mass index.

| Clinical Diagnosis at baseline | Plasma p-tau217 and NfL at baseline | BMI, Underweight | | | BMI, Normal | | | BMI, Overweight | | | BMI, Obese | | |
|---|---|---|---|---|---|---|---|---|---|---|---|---|---|
| | | N | Risk (95% CI) | | N | Risk (95% CI) | | N | Risk (95% CI) | | N | Risk (95% CI) | |
| CU | < cut-off | 6 | - | - | 446 | 2.3 | (1.1–4.6) | 771 | 5.3 | (3.8–7.3) | 377 | 4.2 | (2.4–7.1) |
| | ≥ cut-off | 0 | - | - | 41 | 22.9 | (11.8–39.7) | 44 | 31.7 | (18.7–48.3) | 25 | 27.7 | (12.1–51.4) |
| | All | 14 | 12.4 | (2.6–42.9) | 937 | 7.0 | (5.4–9.1) | 1512 | 8.5 | (7.1–10.2) | 678 | 8.6 | (6.5–11.3) |
| MCI | < cut-off | 2 | - | - | 181 | 18.3 | (12.7–25.6) | 380 | 18 | (13.9–23) | 222 | 25.6 | (19.9–32.3) |
| | ≥ cut-off | 0 | - | - | 29 | 74.0 | (54.9–86.9) | 45 | 57.7 | (41.6–72.3) | 14 | 57.2 | (28.3–81.9) |
| | All | 13 | 21.6 | (5.7–55.6) | 491 | 32.4 | (27.9–37.2) | 801 | 26.3 | (23–30) | 439 | 30.4 | (25.9–35.4) |
| Total | < cut-off | 8 | - | - | 627 | 7.4 | (5.3–10.3) | 1151 | 10.1 | (8.2–12.4) | 599 | 12.8 | (10.1–16.1) |
| | ≥ cut-off | 0 | - | - | 70 | 45.4 | (33.2–58.2) | 89 | 46.6 | (35.6–57.9) | 39 | 39.2 | (24–56.8) |
| | All | 27 | 17.1 | (6.6–37.5) | 1428 | 17.0 | (14.8–19.4) | 2315 | 15.6 | (13.9–17.4) | 1117 | 18.0 | (15.6–20.7) |

p-tau217, phosphorylated tau at threonine 217; NfL, neurofilament light; BMI, body mass index; CI, confidence interval; CU, cognitively unimpaired; MCI, mild cognitive impairment; Two participants had cognitive impairment other than MCI. Plasma p-tau217 data was missing from one participant. Plasma NfL data were missing from 63 participants. BMI data was missing from 29 CU participants and 55 MCI participants. Estimates are weighted.

Table S20. Association between joint intermediate p-tau217 and high NfL range and risk of all-cause dementia at follow-up across demographic and clinical subgroups (N, 2,474).

| Characteristics at baseline | CU | | MCI | | Total | |
|---|---|---|---|---|---|---|
| | RR (95% CI) | | RR (95% CI) | | RR (95% CI) | |
| **All** | 2.39 | (1.19 - 4.81) | 1.36 | (0.76 - 2.45) | 1.63 | (0.99 - 2.71) |
| **Sex** | | | | | | |
| *Female* | 3.52 | (1.70 - 7.29) | 0.84 | (0.32 - 2.17) | 1.55 | (0.84 - 2.83) |
| *Male* | 0.98 | (0.12 - 7.87) | 2.00 | (1.07 - 3.72) | 1.74 | (0.91 - 3.31) |
| **Age** | | | | | | |
| *70–74* | 3.36 | (1.31 - 8.60) | 2.23 | (1.01 - 4.92) | 2.52 | (1.18 - 5.38) |
| *75–79* | 1.08 | (0.26 - 4.46) | 0.58 | (0.15 - 2.30) | 0.71 | (0.19 - 2.71) |
| *80–84* | 2.78 | (1.18 - 6.53) | 1.47 | (0.76 - 2.85) | 1.84 | (0.97 - 3.47) |
| *85–89* | 2.83 | (1.34 - 5.99) | 1.00 | (0.45 - 2.21) | 1.58 | (0.84 - 2.97) |
| *90+* | - | - | - | - | - | - |
| ***APOE* ε4 allele** | | | | | | |
| *ε3ε3/ε3ε2/ε2ε2* | 2.90 | (1.43 - 5.89) | 1.34 | (0.70 - 2.55) | 1.74 | (1.02 - 2.98) |
| *ε2ε4/ε3ε4* | - | - | 1.42 | (0.54 - 3.71 | 1.02 | (0.40 - 2.61) |
| *ε4ε4* | - | - | - | - | - | - |
| **Education** | | | | | | |
| *Primary* | 2.61 | (1.37 - 4.94) | 1.22 | (0.69 - 2.16) | 1.54 | (0.97 - 2.43) |
| *Secondary* | 2.26 | (1.04 - 4.91) | 1.50 | 0.81 - 2.77) | 1.71 | (0.78 - 2.99) |
| *Tertiary* | 2.12 | (0.91 - 4.94) | 1.60 | (0.89 - 2.86) | 1.78 | (1.01 - 3.15) |
| **BMI** | | | | | | |
| *Underweight* | - | - | - | - | - | - |
| *Normal* | 2.54 | 1.30 - 4.97) | 1.06 | (0.56 - 2.02) | 1.33 | (0.77 - 2.30) |
| *Overweight* | 2.16 | (1.03 - 4.54) | 1.42 | (0.84 - 2.42) | 1.66 | (1.01 - 2.73) |
| *Obese* | 2.83 | (1.47 - 5.45) | 1.48 | (0.79 - 2.79) | 1.77 | (1.05 - 2.99) |

p-tau217, phosphorylated tau at threonine 217; NfL, neurofilament light; CU, cognitively unimpaired; MCI, mild cognitive impairment; RR, risk ratio; CI, confidence interval; *APOE*, apolipoprotein E; BMI, body mass index. RRs are given using the low p-tau217 and NfL ranges as reference. Estimates were obtained from a quasi-Poisson regression model with a log link, adjusted for age, sex, education, *APOE* genotype, and BMI, incorporating interaction terms between p-tau217 and NfL ranges, cognitive status and characteristics at baseline. No significant differences between sexes were found when evaluated by rate of risk ratios tests. Inverse probability weighting was applied to reduce bias due to selective participation.

Plasma p-tau217 data was missing from one participant. Plasma NfL data were missing from 63 participants. Two participants had cognitive impairment other than MCI.

#### Supplementary table on biomarkers, comorbidities and risk of all-cause dementia

Table S21. Association between concentrations of plasma p-tau217 and NfL and risk of all-cause dementia according to baseline cognitive status and major comorbidities.

| | Plasma p-tau217 Intermediate range | | | Plasma p-tau217 High range | | | Plasma NfL | | | Plasma p-tau217 + NfL | | |
|---|---|---|---|---|---|---|---|---|---|---|---|---|
| | No self-reported | Self-reported | p-value | No self-reported | Self-reported | p-value | No self-reported | Self-reported | p-value | No self-reported | Self-reported | p-value |
| **Cognitive Unimpaired** | | | | | | | | | | | | |
| *Stroke/brain hemorrhage* | 1.87 (1.22 - 2.86) | 0.81 (0.21 - 3.09) | 0.2124 | 3.40 (2.31- 4.99) | 1.27 (0.46 - 3.53) | 0.0580 | 1.66 (1.10 - 2.49) | 1.52 (0.48 - 4.79) | 0.7885 | 4.12 (1.92 - 8.85) | 1.98 (0.43 - 9.11) | 0.4420 |
| *COPD/emphysema* | 1.87 (1.23 - 2.84) | 0.27 (0.03 - 2.25) | 0.0959 | 3.54 (2.42 - 5.18) | 1.86 (0.69 - 5.07) | 0.4839 | 1.74 (1.17 - 2.58) | 0.53 (0.06 - 4.33) | 0.2689 | 4.66 (2.20 - 9.87) | 1.00 (0.09 - 10.8) | 0.2831 |
| *Cardiovascular disease* | 1.91 (1.25 - 2.93) | 0.75 (0.24 - 2.32) | 0.0970 | 3.45 (2.33 - 5.11) | 1.94 (0.93 - 4.02) | 0.2254 | 1.84 (1.22 - 2.78) | 1.71 (0.75 - 3.92) | 0.7686 | 4.41 (2.06 - 9.44) | 2.95 (0.83 - 10.5) | 0.6110 |
| *Diabetes mellitus* | 1.79 (1.17 - 2.76) | 1.11 (0.48 - 2.53) | 0.1824 | 3.98 (2.71 -5.83) | 0.88 (0.41 -1.87) | **<.0001** | 1.84 (1.25 - 2.69) | 1.14 (0.42 - 3.08) | 0.3842 | 4.96 (2.35 - 10.5) | 0.78 (0.20 - 3.06) | **0.0076** |
| *Rheumatoid arthritis/spondylarthritis* | 1.80 (1.51 - 2.81) | 1.63 (0.65 - 4.04) | 0.5969 | 3.32 (2.24 - 4.93) | 2.88 (1.29 - 6.41) | 0.5380 | 1.88 (1.24 - 2.84) | 1.32 (0.58 - 3.02) | 0.5208 | 4.66 (2.17 - 10.0) | 2.81 (0.82 - 9.85) | 0.1830 |
| **Mild Cognitive Impairment** | | | | | | | | | | | | |
| *Stroke/brain hemorrhage* | 1.25 (0.94 - 1.66) | 1.12 (0.59 - 2.12) | 0.8371 | 1.93 (1.51 - 2.54) | 1.61 (0.88 - 2.95) | 0.8318 | 1.49 (1.06 - 2.11) | 2.67 (1.38 - 5.16) | 0.0608 | 3.20 (2.27 - 4.51) | 4.27 (2.22 - 8.20) | 0.4393 |
| *COPD/emphysema* | 1.22 (0.93 - 1.60) | 1.12 (0.55 - 2.28) | 0.9973 | 1.99 (1.53 - 2.52) | 1.17 (0.62 - 2.21) | 0.1898 | 1.65 (1.10 - 2.47) | 1.61 (0.66 - 3.94) | 0.8988 | 3.72 (2.69 - 5.15) | 2.25 (1.00 - 5.05) | 0.2041 |
| *Cardiovascular disease* | 1.18 (0.89 - 1.57) | 1.32 (0.73 - 2.38) | 0.7689 | 2.03 (1.60 - 2.58) | 1.17 (0.68 - 2.03) | 0.1740 | 1.66 (1.10 - 2.50) | 1.14 (0.65 - 2.00) | 0.4323 | 3.88 (2.80 - 5.39) | 1.60 (0.74 - 3.45) | 0.0941 |
| *Diabetes mellitus* | 1.09 (0.83 - 1.44) | 1.22 (0.77 - 1.93) | 0.3731 | 1.93 (1.53 - 2.42) | 1.46 (0.98 - 2.26) | 0.7377 | 1.64 (1.09 - 2.47) | 1.49 (0.95 - 2.35) | 0.8925 | 3.64 (2.65 - 5.00) | 2.61 (1.56 - 4.37) | 0.2903 |
| *Rheumatoid arthritis/spondylarthritis* | 1.27 (0.96 - 1.67) | 1.05 (0.529 - 2.08) | 0.6759 | 1.91 (1.50 - 2.44) | 1.75 (1.01 - 3.04) | 0.7831 | 1.60 (1.04 - 2.46) | 1.49 (0.86 - 2.57) | 0.8180 | 3.31 (2.27 - 4.82) | 5.01 (2.82 - 8.92) | 0.3536 |
| **Total** | | | | | | | | | | | | |
| *Stroke/brain hemorrhage* | 1.40 (1.04 - 1.77) | 1.04 (0.59 - 1.84) | 0.2277 | 2.28 (1.85 - 2.80) | 1.52 (0.91 - 2.55) | 0.0835 | 1.54 (1.20 - 1.98) | 2.36 (1.33 - 4.20) | 0.6342 | 3.42 (2.46 - 4.76) | 3.60 (2.02 - 6.42) | 0.6984 |
| *COPD/emphysema* | 1.37 (1.09 - 1.72) | 0.88 (0.46 - 1.68) | 0.1182 | 2.35 (1.92 - 2.87) | 1.37 (0.79 - 2.36) | 0.1987 | 1.67 (1.26 - 2.23) | 1.30 (0.57 - 2.96) | 0.2812 | 3.94 (2.88 - 5.38) | 1.86 (0.86 - 4.02) | 0.1453 |
| *Cardiovascular disease* | 1.35 (1.07 - 1.71) | 1.16 (0.70 - 1.93) | 0.1866 | 2.35 (1.92 -2.89) | 1.38 (0.89 - 2.16) | 0.0769 | 1.71 (1.28 - 2.29) | 1.31 (0.83 - 2.08) | 0.5025 | 4.01 (2.93 - 5.48) | 1.91 (0.97 - 3.78) | 0.1924 |
| *Diabetes mellitus* | 1.25 (0.99 - 1.58) | 1.19 (0.80 - 1.77) | 0.4739 | 2.39 (1.96 - 2.91) | 1.32 (0.92 - 1.90) | **0.0001** | 1.70 (0.12 - 2.26) | 1.40 (0.94 - 2.10) | 0.3968 | 3.93 (2.89 - 5.36) | 2.06 (1.29 - 3.27) | **0.0041** |
| *Rheumatoid arthritis/spondylarthritis* | 1.39 (1.10 - 1.76) | 1.21 (0.70 - 2.09) | 0.5009 | 2.24 (1.82 - 2.76) | 2.06 (1.30 - 3.25) | 0.5067 | 1.68 (1.24 - 2.26) | 1.43 (0.91 - 2.25) | 0.6785 | 3.62 (2.56 - 5.14) | 4.35 (2.60 - 7.28) | 0.7660 |

p-tau217, phosphorylated tau at threonine 217; NfL, neurofilament light. Weighted risk ratios with 95% confidence intervals for all-cause dementia according to plasma p-tau217 range, plasma NfL range, and p-tau217 and NfL combination, stratified by baseline cognitive status and major comorbidities. Estimates are derived from survey-weighted quasi-Poisson regression models adjusted for age, sex, education, APOE genotype, and serum creatinine. Low range p-tau217 served as the reference category. Results are shown separately for participants with and without each self-reported comorbidity. *P* values correspond to interaction tests between p-tau217 range and comorbidity status within cognitive strata.

#### Supplementary tables on p-tau217 and AD dementia

Table S22. Estimated absolute risk of clinical Alzheimer's disease dementia by baseline cognitive status, plasma p-tau217 range and sex.

| Clinical Diagnosis at baseline | Plasma p-tau217 at baseline | Female | | | Male | | | Total | | |
|---|---|---|---|---|---|---|---|---|---|---|
| | | N | Risk (95% CI) | | N | Risk (95% CI) | | N | Risk (95% CI) | |
| CU | < 0.40 | 995 | 3.7 | (2.6–5.3) | 721 | 3.1 | (2–4.9) | 1716 | 3.5 | (2.6–4.6) |
| | 0.40–0.63 | 428 | 6.4 | (4.3–9.6) | 383 | 5.9 | (3.8–9.1) | 811 | 6.2 | (4.6–8.3) |
| | ≥ 0.63 | 323 | 14.6 | (10.9–19.3) | 320 | 9.9 | (6.8–14.3) | 643 | 12.4 | (9.8–15.4) |
| | All | 1746 | 6.6 | (5.4–8.0) | 1424 | 5.5 | (4.3–7.0) | 3170 | 6.1 | (5.2–7.1) |
| MCI | < 0.40 | 435 | 16.1 | (12.4–20.6) | 420 | 11.1 | (8.2–14.9) | 855 | 13.8 | (11.3–16.7) |
| | 0.40–0.63 | 229 | 22.5 | (16.8–29.4) | 258 | 17.8 | (13–23.9) | 487 | 20.1 | (16.2–24.5) |
| | ≥ 0.63 | 230 | 42.2 | (35.3–49.3) | 227 | 32.3 | (26.1–39.2) | 457 | 37.6 | (32.9–42.6) |
| | All | 894 | 25.4 | (22.2–28.8) | 905 | 18.7 | (16.1–21.8) | 1799 | 22.2 | (20.1–24.5) |
| Total | < 0.40 | 1430 | 8.0 | (6.5–9.9) | 1141 | 6.4 | (5.0–8.2) | 2571 | 7.3 | (6.2–8.6) |
| | 0.40–0.63 | 657 | 12.8 | (10–16.2) | 641 | 11.2 | (8.6–14.4) | 1298 | 12.1 | (10.1–14.3) |
| | ≥ 0.63 | 554 | 28.1 | (24–32.6) | 547 | 20.3 | (16.8–24.3) | 1101 | 24.4 | (21.7–27.5) |
| | All | 2641 | 14.0 | (12.5–15.7) | 2329 | 11.2 | (9.8–12.8) | 4970 | 12.7 | (11.7–13.9) |

p-tau217, phosphorylated tau at threonine 217; CI, confidence interval; CU, cognitively unimpaired; MCI, mild cognitive impairment; Concentration is given in picograms per milliliter. Plasma p-tau217 data was missing from one participant. Two participants had cognitive impairment other than MCI. Estimates are weighted.

Table S23. Estimated absolute risk of clinical Alzheimer's disease dementia by baseline cognitive status, plasma p-tau217 range and age groups.

| Clinical Diagnosis at baseline | Plasma p-tau217 at baseline | Age, 70–74 | | | Age, 75–79 | | | Age, 80–84 | | | Age, 85–89 | | | Age, 90+ | | |
|---|---|---|---|---|---|---|---|---|---|---|---|---|---|---|---|---|
| | | N | Risk (95% CI) | | N | Risk (95% CI) | | N | Risk (95% CI) | | N | Risk (95% CI) | | N | Risk (95% CI) | |
| CU | < 0.40 | 1038 | 2.5 | (1.6–3.8) | 472 | 3.2 | (1.9–5.6) | 163 | 5.4 | (2.8–10.3) | 32 | 11.1 | (3.9–28) | 11 | 21.4 | (5.6–55.5) |
| | 0.40–0.63 | 377 | 3.8 | (2.2–6.3) | 268 | 6.0 | (3.6–9.9) | 122 | 8.8 | (4.7–15.9) | 40 | 15.0 | (6.2–32.3) | 4 | - | - |
| | ≥ 0.63 | 220 | 6.8 | (4–11.4) | 216 | 9.2 | (5.8–14.1) | 141 | 17.4 | (11.8–25.1) | 52 | 22.2 | (12.4–36.7) | 14 | 23.2 | (6.6–56.5) |
| | All | 1635 | 3.4 | (2.5–4.5) | 956 | 5.4 | (4–7.1) | 426 | 10.5 | (7.8–14) | 124 | 16.8 | (10.9–25) | 29 | 18.8 | (8.1–37.9) |
| MCI | < 0.40 | 519 | 7.6 | (5.5–10.6) | 246 | 19.3 | (14.5–25.4) | 73 | 16.5 | (9.3–27.8) | 12 | 53.1 | (23.2–80.9) | 5 | 53.9 | (7.9–94.1) |
| | 0.40–0.63 | 237 | 11.8 | (8–17) | 149 | 19.6 | (13.5–27.6) | 68 | 27.1 | (17–40.3) | 28 | 38.2 | (21.3–58.6) | 5 | 45.6 | (5.9–91.9) |
| | ≥ 0.63 | 180 | 27.1 | (20.8–34.6) | 136 | 29.1 | (21.8–37.7) | 85 | 50.7 | (39.5–61.8) | 44 | 48.3 | (33–63.9) | 12 | 74.3 | (39.1–92.8) |
| | All | 936 | 12.6 | (10.5–15.1) | 531 | 22.0 | (18.4–26) | 226 | 33.0 | (26.7–40) | 84 | 45.6 | (34.7–57) | 22 | 61.2 | (37.7–80.5) |
| Total | < 0.40 | 1557 | 4.4 | (3.4–5.7) | 718 | 9.4 | (7.2–12.2) | 236 | 9.4 | (6.1–14.3) | 44 | 25.8 | (13.8–42.9) | 16 | 35.4 | (14.2–64.5) |
| | 0.40–0.63 | 614 | 7.1 | (5.2–9.7) | 417 | 11.4 | (8.4–15.2) | 190 | 16.4 | (11.2–23.3) | 68 | 26.8 | (16.8–40) | 9 | 29.7 | (6–73.6) |
| | ≥ 0.63 | 401 | 17.1 | (13.4–21.5) | 352 | 17.9 | (14–22.6) | 226 | 32.0 | (25.7–39.1) | 96 | 37 | (27–48.1) | 26 | 50.1 | (30.1–70) |
| | All | 2572 | 7.1 | (6.1–8.3) | 1487 | 12.0 | (10.3–14) | 652 | 19.6 | (16.4–23.3) | 208 | 31.2 | (24.7–38.5) | 51 | 41.0 | (27.4–56.2) |

p-tau217, phosphorylated tau at threonine 217; CI, confidence interval; CU, cognitively unimpaired; MCI, mild cognitive impairment; Concentration is given in picograms per milliliter. Age is given in years. Plasma p-tau217 data was missing from one participant. Two participants had cognitive impairment other than MCI. Estimates are weighted.

Table S24. Estimated absolute risk of clinical Alzheimer's disease dementia by baseline cognitive status, plasma p-tau217 range and *APOE* genotype.

| **Clinical Diagnosis at baseline** | **Plasma p-tau217 at baseline** | **ε3ε3/ε3ε2/ε2ε2** | | | **ε2ε4/ε3ε4** | | | **ε4ε4** | | |
|---|---|---|---|---|---|---|---|---|---|---|
| | | **N** | **Risk** | **(95% CI)** | **N** | **Risk** | **(95% CI)** | **N** | **Risk** | **(95% CI)** |
| CU | < 0.40 | 1391 | 3.7 | (2.7–4.9) | 305 | 2.9 | (1.4–5.8) | 14 | - | - |
| | 0.40–0.63 | 528 | 6.4 | (4.4–9.3) | 266 | 5.2 | (3–8.9) | 14 | 6.9 | (0.8–41.1) |
| | ≥ 0.63 | 374 | 10.4 | (7.4–14.4) | 240 | 13.6 | (9.5–19.3) | 27 | 30.2 | (15.1–51.3) |
| | All | 2293 | 5.5 | (4.5–6.7) | 811 | 7.0 | (5.2–9.2) | 55 | 16.5 | (8.6–29.5) |
| MCI | < 0.40 | 694 | 14.5 | (11.7–17.7) | 154 | 10.1 | (5.7–17.2) | 2 | - | - |
| | 0.40–0.63 | 308 | 20.7 | (16–26.3) | 157 | 19.2 | (12.7–27.9) | 16 | 25.1 | (8.8–53.6) |
| | ≥ 0.63 | 224 | 36.5 | (29.8–43.7) | 192 | 37.7 | (30.6–45.4) | 36 | 43.4 | (26.6–61.8) |
| | All | 1226 | 20.8 | (18.3–23.5) | 503 | 23.9 | (19.9–28.4) | 54 | 36.9 | (23.9–52.1) |
| Total | < 0.40 | 2085 | 7.7 | (6.4–9.2) | 459 | 5.6 | (3.6–8.7) | 16 | - | - |
| | 0.40–0.63 | 836 | 12.3 | (9.9–15.2) | 423 | 11.2 | (7.9–15.5) | 30 | 17.0 | (6.9–36) |
| | ≥ 0.63 | 598 | 21.9 | (18.3–26) | 433 | 25.8 | (21.4–30.7) | 63 | 38.3 | (26–52.3) |
| | All | 3520 | 11.5 | (10.3–12.9) | 1315 | 14.4 | (12.3–16.8) | 109 | 27.3 | (19.0–37.6) |

p-tau217, phosphorylated tau at threonine 217; *APOE*, apolipoprotein E; CI, confidence interval; CU, cognitively unimpaired; MCI, mild cognitive impairment; Concentration is given in picograms per milliliter. Plasma p-tau217 data was missing from one participant. *APOE* allele status was missing for 11 CU and 16 MCI participants. Two participants had cognitive impairment other than MCI. Estimates are weighted.

Table S25. Estimated absolute risk of clinical Alzheimer's disease dementia by baseline cognitive status, plasma p-tau217 range and educational level.

| Clinical Diagnosis at baseline | Plasma p-tau217 at baseline | Primary education | | | Secondary education | | | Tertiary education | | |
|---|---|---|---|---|---|---|---|---|---|---|
| | | N | Risk | (95% CI) | N | Risk | (95% CI) | N | Risk | (95% CI) |
| CU | < 0.40 | 326 | 4.9 | (2.9–8) | 784 | 3.7 | (2.5–5.3) | 606 | 1.7 | (0.9–3.2) |
| | 0.40–0.63 | 169 | 7.0 | (3.9–12.3) | 373 | 7.1 | (4.8–10.4) | 268 | 3.0 | (1.5–6) |
| | ≥ 0.63 | 151 | 14.1 | (9.2–21) | 269 | 12.3 | (8.7–17.1) | 223 | 10.1 | (6.6–15) |
| | All | 646 | 7.7 | (5.8–10.2) | 1426 | 6.3 | (5.1–7.8) | 1097 | 3.8 | (2.8–5.2) |
| MCI | < 0.40 | 220 | 19.8 | (14.7–26.2) | 416 | 12.4 | (9.4–16) | 219 | 4.7 | (2.4–8.8) |
| | 0.40–0.63 | 115 | 29.6 | (21.5–39.2) | 239 | 15.1 | (10.9–20.7) | 131 | 12.8 | (7.8–20.3) |
| | ≥ 0.63 | 146 | 44.5 | (36.3–53.1) | 208 | 33.8 | (27.5–40.9) | 102 | 27.9 | (19.9–37.6) |
| | All | 481 | 30.2 | (26.0–34.8) | 863 | 18.7 | (16.1–21.7) | 452 | 12.5 | (9.6–16.1) |
| Total | < 0.40 | 546 | 11.5 | (8.8–14.8) | 1200 | 6.9 | (5.5–8.6) | 825 | 2.5 | (1.6–4) |
| | 0.40–0.63 | 284 | 17.4 | (13.1–22.9) | 612 | 10.5 | (8.1–13.4) | 399 | 6.4 | (4.3–9.6) |
| | ≥ 0.63 | 297 | 30.7 | (25.4–36.7) | 477 | 22.5 | (18.8–26.8) | 326 | 16.3 | (12.5–20.9) |
| | All | 1127 | 18.5 | (16.1–21.1) | 2290 | 11.4 | (10.1–12.9) | 1550 | 6.6 | (5.4–8.0) |

p-tau217, phosphorylated tau at threonine 217; CI, confidence interval; CU, cognitively unimpaired; MCI, mild cognitive impairment; Concentration is given in picograms per milliliter. Plasma p-tau217 data was missing from one participant. Education status was missing for four participants. Two participants had cognitive impairment other than MCI. Estimates are weighted. "Primary education" was defined ≤10 years of compulsory primary and lower secondary education; "Secondary education" was defined as additional 1–2 years of academic or vocational school, 3 years of academic or vocational school, or 3–4 years of vocational training or apprenticeship (upper secondary education); "Tertiary education" referred to college or university education.

Table S26. Estimated absolute risk of clinical Alzheimer's disease dementia by baseline cognitive status, plasma p-tau217 range and body mass index.

| Clinical Diagnosis | Plasma p-tau217 | BMI, Underweight | | | BMI, Normal | | | BMI, Overweight | | | BMI, Obese | | |
|---|---|---|---|---|---|---|---|---|---|---|---|---|---|
| at baseline | at baseline | N | Risk | (95% CI) | N | Risk | (95% CI) | N | Risk | (95% CI) | N | Risk | (95% CI) |
| CU | < 0.40 | 7 | 14.2 | (1.2–70.2) | 476 | 2.5 | (1.2–4.8) | 826 | 3.8 | (2.6–5.5) | 392 | 2.4 | (1.2–4.7) |
| | 0.40–0.63 | 2 | - | - | 245 | 6.2 | (3.7–10.5) | 400 | 5.7 | (3.6–8.9) | 160 | 7.6 | (4–14) |
| | ≥ 0.63 | 5 | 15.3 | (0.8–80.3) | 216 | 11.3 | (7.5–16.8) | 286 | 14.2 | (10.3–19.4) | 126 | 9.6 | (5.2–17) |
| | All | 14 | 12.4 | (2.6–42.9) | 937 | 5.6 | (4.2–7.5) | 1512 | 6.4 | (5.2–8) | 678 | 5.1 | (3.5–7.3) |
| MCI | < 0.40 | 2 | - | - | 202 | 11.6 | (7.4–17.8) | 410 | 12.1 | (8.8–16.5) | 232 | 17.2 | (12.5–23.2) |
| | 0.40–0.63 | 6 | 33.1 | (4.6–83.5) | 135 | 13.0 | (7.9–20.7) | 208 | 20.1 | (14.4–27.4) | 114 | 22.5 | (14.8–32.6) |
| | ≥ 0.63 | 5 | 15.3 | (0.8–80.6) | 154 | 47.1 | (38.7–55.7) | 183 | 31.2 | (24.2–39.1) | 93 | 26.7 | (18.2–37.4) |
| | All | 13 | 21.6 | (5.7–55.6) | 491 | 24.3 | (20.3–29) | 801 | 19 | (16–22.4) | 439 | 20.8 | (16.9–25.4) |
| Total | < 0.40 | 9 | 10.8 | (1–58.5) | 678 | 5.6 | (3.8–8.1) | 1236 | 7.0 | (5.4–8.9) | 624 | 8.3 | (6.2–11.1) |
| | 0.40–0.63 | 8 | 25.3 | (4.3–72) | 380 | 8.9 | (6.2–12.7) | 608 | 11.2 | (8.5–14.6) | 274 | 14.4 | (10.1–20.1) |
| | ≥ 0.63 | 10 | 15.3 | (2.9–52.3) | 370 | 28.4 | (23.4–33.9) | 470 | 21.9 | (18–26.5) | 219 | 17.7 | (12.8–24) |
| | All | 27 | 17.1 | (6.6–37.5) | 1428 | 13.0 | (11–15.2) | 2315 | 11.4 | (10–13.1) | 1117 | 11.8 | (9.8–14.2) |

p-tau217, phosphorylated tau at threonine 217; BMI, body mass index; CI, confidence interval; CU, cognitively unimpaired; MCI, mild cognitive impairment; Concentration is given in picograms per milliliter. Plasma p-tau217 data was missing from one participant. BMI data was missing for 29 CU participants and 55 MCI participants. Two participants had cognitive impairment other than MCI. Estimates are weighted.

Table S27. Association between plasma p-tau217 range at baseline and risk of clinical Alzheimer's disease dementia at follow-up across demographic and clinical subgroups (N=4,857).

| Characteristics | CU | MCI | Total | CU | MCI | Total |
|---|---|---|---|---|---|---|
| at baseline | RR (95% CI) | RR (95% CI) | RR (95% CI) | RR (95% CI) | RR (95% CI) | RR (95% CI) |
| | *Intermediate range of plasma p-tau217* | | | *High range of plasma p-tau217* | | |
| **All** | 1.64 (1.06–2.54) | 1.24 (0.92–1.67) | 1.34 (1.05–1.72) | 2.74 (1.85–4.07) | 1.94 (1.50–2.52) | 2.15 (1.72–2.67) |
| **Sex** | | | | | | |
| *Female* | 1.80 (1.03–3.15) | 1.15 (0.77–1.73) | 1.32 (0.95–1.82) | 3.31 (2.02–5.42) | 1.88 (1.37–2.57) | 2.24 (1.71–2.93) |
| *Male* | 1.44 (0.75–2.79) | 1.35 (0.88–2.08) | 1.38 (0.96–1.97) | 2.01 (1.09–3.69) | 2.03 (1.38–2.99) | 2.03 (1.46–2.81) |
| **Age** | | | | | | |
| *70–74* | 1.68 (0.85–3.35) | 1.88 (1.12–3.16) | 1.81 (1.20–2.74) | 2.56 (1.29–5.1) | 3.52 (2.29–5.41) | 3.19 (2.22–4.58) |
| *75–79* | 1.71 (0.78–3.78) | 1.06 (0.65–1.71) | 1.20 (0.79–1.80) | 2.89 (1.41–5.93) | 1.52 (1.01–2.29) | 1.81 (1.27–2.57) |
| *80–84* | 1.66 (0.69–3.99) | 1.70 (0.84–3.42) | 1.68 (0.97–2.91) | 2.98 (1.41–6.30) | 2.73 (1.49–5.00) | 2.82 (1.76–4.52) |
| *85–89* | 1.99 (0.46–8.66) | 0.78 (0.36–1.72) | 0.96 (0.49–1.90) | 2.93 (0.76–11.2) | 0.85 (0.45–1.61) | 1.16 (0.66–2.06) |
| *90+* | - - | 0.63 (0.19–2.09) | 0.49 (0.15–1.55) | 1.21 (0.17–8.82) | 1.19 (0.55–2.61) | 1.20 (0.56–2.56) |
| ***APOE* ε4 allele** | | | | | | |
| *ε3ε3/ε3ε2/ε2ε2* | 1.62 (0.99–2.65) | 1.12 (0.79–1.57) | 1.24 (0.94–1.65) | 2.29 (1.44–3.64) | 1.57 (1.16–2.12) | 1.76 (1.36–2.26) |
| *ε2ε4/ε3ε4* | 1.54 (0.63–3.81) | 1.39 (0.74–2.63) | 1.43 (0.85–2.42) | 3.55 (1.60–7.87) | 2.83 (1.63–4.89) | 3.02 (1.91–4.76) |
| *ε4ε4* | - - | - - | - - | - - | - - | - - |
| **Education** | | | | | | |
| *Primary* | 1.58 (0.71–3.52) | 1.24 (0.81–1.90) | 1.30 (0.89–1.90) | 2.77 (1.38–5.56) | 1.60 (1.12–2.29) | 1.82 (1.32–2.51) |
| *Secondary* | 1.72 (0.98–3.01) | 1.02 (0.66–1.6) | 1.24 (0.87–1.75) | 2.27 (1.33–3.86) | 2.01 (1.41–2.88) | 2.09 (1.55–2.82) |
| *Tertiary* | 1.55 (0.59–4.06) | 2.66 (1.22–5.82) | 2.16 (1.19–3.92) | 4.36 (1.98–9.59) | 4.76 (2.34–9.67) | 4.58 (2.70–7.75) |
| **BMI** | | | | | | |
| *Underweight* | - - | - - | 2.86 (0.30–26.9) | 1.28 (0.09–18.8) | - - | 2.70 (0.25–29.3) |
| *Normal* | 2.15 (0.87–5.29) | 1.10 (0.59–2.08) | 1.36 (0.82–2.27) | 3.41 (1.49–7.78) | 3.21 (2.04–5.02) | 3.25 (2.19–4.83) |
| *Overweight* | 1.14 (0.64–2.05) | 1.45 (0.93–2.28) | 1.35 (0.95–1.94) | 2.44 (1.47–4.07) | 1.91 (1.28–2.84) | 2.08 (1.52–2.86) |
| *Obese* | 2.89 (1.18–7.12) | 1.02 (0.64–1.63) | 1.29 (0.85–1.96) | 3.03 (1.28–7.17) | 1.08 (0.66–1.76) | 1.36 (0.90–2.07) |

p-tau217, phosphorylated tau at threonine 217; CU, cognitively unimpaired; MCI, mild cognitive impairment; RR, risk ratio; CI, confidence interval; *APOE*, apolipoprotein E; BMI, body mass index. RRs are given using the low p-tau217 range as reference. Estimates were obtained from a quasi-Poisson regression model with a log link, adjusted for age, sex, education, *APOE* genotype, and BMI, incorporating interaction terms between p-tau217 range, cognitive status and characteristics at HUNT4 70+. No significant differences between sexes were found when evaluated by rate of risk ratios tests. Inverse probability weighting was applied to reduce bias due to selective participation. Plasma p-tau217 data was missing from one participant. Two participants had cognitive impairment other than MCI.

#### Supplementary tables on NfL and AD dementia

Table S28. Estimated absolute risk of clinical Alzheimer's disease dementia by baseline cognitive status, plasma NfL range and sex.

| Clinical Diagnosis at baseline | Plasma NfL at baseline | Female | | | Male | | | Total | | |
|---|---|---|---|---|---|---|---|---|---|---|
| | | N | Risk (95% CI) | | N | Risk (95% CI) | | N | Risk (95% CI) | |
| CU | < cut-off | 1583 | 6.2 | (5.0–7.6) | 1295 | 4.5 | (3.4–5.9) | 2878 | 5.4 | (4.6–6.4) |
| | ≥ cut-off | 140 | 11.1 | (6.5–18.4) | 119 | 16.3 | (10.0–25.4) | 259 | 13.4 | (9.4–18.8) |
| | All | 1746 | 6.6 | (5.4–8.0) | 1424 | 5.5 | (4.3–7.0) | 3170 | 6.1 | (5.2–7.1) |
| MCI | < cut-off | 779 | 23.6 | (20.3–27.3) | 792 | 17.5 | (14.7–20.6) | 1571 | 20.6 | (18.4–23.0) |
| | ≥ cut-off | 103 | 36.5 | (26.9–47.2) | 95 | 29.3 | (20.0–40.8) | 198 | 33.4 | (26.5–41.1) |
| | All | 894 | 25.4 | (22.2–28.8) | 905 | 18.7 | (16.1–21.8) | 1799 | 22.2 | (20.1–24.5) |
| Total | < cut-off | 2364 | 12.8 | (11.3–14.5) | 2087 | 10.0 | (8.6–11.5) | 4451 | 11.5 | (10.4–12.7) |
| | ≥ cut-off | 243 | 23.9 | (18.2–30.6) | 214 | 22.7 | (16.9–29.9) | 457 | 23.4 | (19.2–28.2) |
| | All | 2642 | 14.0 | (12.5–15.7) | 2329 | 11.2 | (9.8–12.8) | 4971 | 12.7 | (11.7–13.9) |

NfL, neurofilament light; CI, confidence interval; CU, cognitively unimpaired; MCI, mild cognitive impairment; Plasma NfL data were missing from 35 females and 28 males. Two participants had cognitive impairment other than MCI. Estimates are weighted.

Table S29. Estimated absolute risk of clinical Alzheimer's disease dementia by baseline cognitive status, plasma NfL range and age groups.

| Clinical Diagnosis at baseline | Plasma NfL at baseline | Age, 70–74 | | | Age, 75–79 | | | Age, 80–84 | | | Age, 85–89 | | | Age, 90+ | | |
|---|---|---|---|---|---|---|---|---|---|---|---|---|---|---|---|---|
| | | N | Risk (95% CI) | | N | Risk (95% CI) | | N | Risk (95% CI) | | N | Risk (95% CI) | | N | Risk (95% CI) | |
| CU | < cut-off | 1501 | 3.0 | (2.2–4.1) | 897 | 5.1 | (3.8–6.9) | 375 | 10.1 | (7.3–13.8) | 93 | 13.7 | (7.8–23.0) | 14 | 19.0 | (5.3–49.7) |
| | ≥ cut-off | 111 | 8.0 | (3.8–16.3) | 53 | 9.7 | (4.0–21.7) | 50 | 14.0 | (6.5–27.6) | 31 | 25.4 | (12.3–45.2) | 14 | 20.9 | (5.7–53.3) |
| | All | 1635 | 3.4 | (2.5–4.5) | 956 | 5.4 | (4.0–7.1) | 426 | 10.5 | (7.8–14.0) | 124 | 16.8 | (10.9–25.0) | 29 | 18.8 | 8.1–37.9) |
| MCI | < cut-off | 841 | 11.2 | (9.1–13.7) | 483 | 22.6 | (18.9–26.9) | 181 | 31.0 | (24.1–38.8) | 56 | 49.4 | (35.8–63.1) | 10 | 71.1 | (32.8–92.5) |
| | ≥ cut-off | 78 | 24.9 | (15.9–36.6) | 39 | 19.8 | (9.5–37.0) | 43 | 41.0 | (26.1–57.6) | 26 | 40.9 | (22.5–62.2) | 12 | 52.5 | (22.6–80.8) |
| | All | 963 | 12.6 | (10.5–15.1) | 531 | 22.0 | (18.4–26.0) | 226 | 33.0 | (26.7–40.0) | 84 | 45.6 | (34.7–57.0) | 22 | 61.2 | (37.7–80.5) |
| Total | < cut-off | 2343 | 6.3 | (5.3 -7.5) | 1381 | 12.0 | (10.2–14.0) | 554 | 18.1 | (14.8–22.0) | 149 | 30.7 | (23.1–39.5) | 24 | 45.2 | (24.9–67.2) |
| | ≥ cut-off | 189 | 15.7 | (10.7–22.5) | 92 | 14.5 | (8.3–24.3) | 93 | 28.0 | (18.9–39.3) | 57 | 33.8 | (21.7–48.3) | 26 | 38.9 | (20.9–60.5) |
| | All | 2572 | 7.1 | (6.1–8.3) | 1488 | 12.0 | (10.3–14.0) | 653 | 19.6 | (16.4–23.3) | 208 | 31.2 | (24.7–38.5) | 51 | 41.0 | (27.4–56.2) |

NfL, neurofilament light; CI, confidence interval; CU, cognitively unimpaired; MCI, mild cognitive impairment. Age is given in years. Plasma NfL data were missing from 63 participants. Two participants had cognitive impairment other than MCI. Estimates are weighted.

Table S30. Estimated absolute risk of clinical Alzheimer's disease dementia by baseline cognitive status, plasma NfL range and *APOE* genotype.

| Clinical Diagnosis at baseline | Plasma NfL at baseline | ε3ε3/ε3ε2/ε2ε2 | | | ε2ε4/ε3ε4 | | | ε4ε4 | | |
|---|---|---|---|---|---|---|---|---|---|---|
| | | N | Risk | (95% CI) | N | Risk | (95% CI) | N | Risk | (95% CI) |
| CU | < cut-off | 2084 | 5.2 | (4.2–6.4) | 733 | 5.4 | (3.9–7.4) | 50 | 13.6 | (6.4–26.7) |
| | ≥ cut-off | 185 | 9.3 | (5.6–14.9) | 69 | 22.3 | (12.8–35.9) | 5 | 49.6 | (7.1–92.7) |
| | All | 2293 | 5.5 | (4.5–6.7) | 811 | 7.0 | (5.2–9.2) | 55 | 16.5 | (8.6–29.5) |
| MCI | < cut-off | 1067 | 19.1 | (16.5–21.9) | 442 | 22.6 | (18.4–27.3) | 47 | 34 | (20.7–50.4) |
| | ≥ cut-off | 135 | 33.2 | (24.9–42.6) | 56 | 34.0 | (21.5–49.3) | 6 | 40.9 | (6.2–87.9) |
| | All | 1226 | 20.8 | (18.3–23.5) | 503 | 23.9 | (19.9–28.4) | 54 | 36.9 | (23.9–52.1) |
| Total | < cut-off | 3152 | 10.5 | (9.3–11.8) | 1176 | 12.9 | (10.8–15.3) | 97 | 24.1 | (15.9–34.7) |
| | ≥ cut-off | 320 | 21.2 | (16.4–26.4) | 125 | 28.0 | (19.9–37.9) | 11 | 44.5 | (15.8–77.4) |
| | All | 3520 | 11.5 | (10.3–12.9) | 1315 | 14.4 | (12.3–16.8) | 109 | 27.3 | (19.0–37.6) |

NfL, neurofilament light; *APOE*, apolipoprotein E; CI, confidence interval; CU, cognitively unimpaired; MCI, mild cognitive impairment; Plasma NfL data were missing from 63 participants. *APOE* genotype was missing for 11 CU and 16 MCI participants. Two participants had cognitive impairment other than MCI. Estimates are weighted.

Table S31. Estimated absolute risk of clinical Alzheimer's disease dementia by baseline cognitive status, plasma NfL range and educational level.

| Clinical Diagnosis at baseline | Plasma NfL at baseline | Primary education | | | Secondary education | | | Tertiary education | | |
|---|---|---|---|---|---|---|---|---|---|---|
| | | N | Risk (95% CI) | | N | Risk (95% CI) | | N | Risk (95% CI) | |
| CU | < cut-off | 557 | 7.0 | (5.1–9.5) | 1300 | 5.8 | (4.6–7.3) | 1001 | 3.1 | (2.1–4.4) |
| | ≥ cut-off | 59 | 15.0 | (7.6–27.7) | 108 | 12.1 | (6.9–20.4) | 91 | 11.7 | (6.2–21.1) |
| | All | 646 | 7.7 | (5.8–10.2) | 1426 | 6.3 | (5.1–7.8) | 1097 | 3.8 | (2.8–5.2) |
| MCI | < cut-off | 417 | 28.7 | (24.3–33.6) | 747 | 17.4 | (14.7–20.5) | 406 | 11.2 | (8.4–14.9) |
| | ≥ cut-off | 57 | 40.9 | (28.3–54.9) | 98 | 28.0 | (19.6–38.4) | 41 | 25.4 | (13.9–41.7) |
| | All | 481 | 30.2 | (26.0–34.8) | 863 | 18.7 | (16.1–21.7) | 452 | 12.5 | (9.6–16.1) |
| Total | < cut-off | 994 | 17.1 | (14.7–19.9) | 2048 | 10.4 | (9.0–11.9) | 1408 | 5.6 | (4.5–7.1) |
| | ≥ cut-off | 116 | 29.4 | (21.2–39.1) | 206 | 20.4 | (15.1–26.9) | 132 | 16.3 | (10.6–24.2) |
| | All | 1127 | 18.5 | (16.1–21.1) | 2290 | 11.4 | (10.1–12.9) | 1550 | 6.6 | (5.4–8.0) |

NfL, neurofilament light; CI, confidence interval; CU, cognitively unimpaired; MCI, mild cognitive impairment; Plasma NfL data were missing from 63 participants. Education status was missing from four participants. Two participants had cognitive impairment other than MCI. Estimates are weighted. "Primary education" was defined ≤10 years of compulsory primary and lower secondary education; "Secondary education" was defined as additional 1–2 years of academic or vocational school, 3 years of academic or vocational school, or 3–4 years of vocational training or apprenticeship (upper secondary education); "Tertiary education" referred to college or university education.

Table S32. Estimated absolute risk of clinical Alzheimer's disease dementia by baseline cognitive status, plasma NfL range and body mass index.

| Clinical Diagnosis at baseline | Plasma NfL at baseline | BMI, Underweight | | | BMI, Normal | | | BMI, Overweight | | | BMI, Obese | | |
|---|---|---|---|---|---|---|---|---|---|---|---|---|---|
| | | N | Risk | (95% CI) | N | Risk | (95% CI) | N | Risk | (95% CI) | N | Risk | (95% CI) |
| CU | < cut-off | 13 | 5.7 | (0.6–36.8) | 840 | 5.1 | (3.7–7) | 1374 | 5.8 | (4.5–7.3) | 629 | 4.4 | (3–6.6) |
| | ≥ cut-off | 1 | 100 | - | 90 | 10.9 | (5.6–20.1) | 117 | 14.2 | (8.4–23) | 44 | 13.2 | (5.1–30.1) |
| | All | 14 | 12.4 | (2.6–42.9) | 937 | 5.6 | (4.2–7.5) | 1512 | 6.4 | (5.2–8) | 678 | 5.1 | (3.5–7.3) |
| MCI | < cut-off | 13 | 21.6 | (5.7–55.6) | 420 | 24.0 | (19.7–29) | 698 | 16.7 | (13.7–20.2) | 409 | 21.1 | (17–25.8) |
| | ≥ cut-off | 0 | | | 62 | 26.2 | (15.6–40.6) | 87 | 36.2 | (25.7–48) | 26 | 16.1 | (5.6–38.4) |
| | All | 13 | 21.6 | (5.7–55.6) | 491 | 24.3 | (20.3–29) | 801 | 19.0 | (16–22.4) | 439 | 20.8 | (16.9–25.4) |
| Total | < cut-off | 26 | 14.1 | (4.8–34.9) | 1260 | 12.3 | (10.3–14.6) | 2074 | 10 | (8.5–11.6) | 1038 | 11.6 | (9.6–14.1) |
| | ≥ cut-off | 1 | 100 | - | 152 | 17.9 | (11.9–26.1) | 204 | 25.1 | (18.9–32.6) | 70 | 14.3 | (7.2–26.4) |
| | All | 27 | 17.1 | (6.6–37.5) | 1428 | 13.0 | (11–15.2) | 2315 | 11.4 | (10–13.1) | 1117 | 11.8 | (9.8–14.2) |

NfL, neurofilament light; BMI, Body mass index; CI, confidence interval; CU, cognitively unimpaired; MCI, mild cognitive impairment; Plasma NfL data were missing from 63 participants. BMI data were missing from 29 CU participants and 55 MCI participants. Two participants had cognitive impairment other than MCI. Estimates are weighted.

Table S33. Association between baseline high range plasma neurofilament light and risk of clinical Alzheimer's disease dementia at follow-up across demographic and clinical subgroups (N, 4,795).

| Characteristics at baseline | CU | MCI | Total |
|---|---|---|---|
| | RR (95% CI) | RR (95% CI) | RR (95% CI) |
| All | 1.86 (1.255–2.76) | 1.58 (0.886–2.83) | 1.66 (1.12–2.46) |
| Sex | | | |
| *Female* | 1.58 (0.945–2.65) | 1.72 (0.726–4.08) | 1.68 (0.918–3.07) |
| *Male* | 2.33 (1.293–4.18) | 1.40 (0.948–2.08) | 1.64 (1.184–2.27) |
| Age | | | |
| *70–74* | 2.60 (1.131–5.96) | 2.94 (1.301–6.64) | 2.84 (1.554–5.19) |
| *75–79* | 1.73 (0.688–4.33) | 1.13 (0.412–3.09) | 1.28 (0.619–2.66) |
| *80–84* | 1.31 (0.605–2.84) | 1.45 (0.85–2.47) | 1.40 (0.907–2.15) |
| *85–89* | 2.15 (0.95–4.87) | 0.83 (0.436–1.58) | 1.15 (0.703–1.88) |
| *90+* | 1.09 (0.175–6.8) | 0.65 (0.275–1.54) | 0.74 (0.342–1.58) |
| *APOE* ε4 allele | | | |
| *ε3ε3/ε3ε2/ε2ε2* | 1.58 (0.941–2.66) | 1.40 (0.815–2.41) | 1.45 (1.002–2.11) |
| *ε2ε4/ε3ε4* | 2.36 (1.225–4.54) | 1.94 (0.883–4.28) | 2.05 (1.136–3.7) |
| *ε4ε4* | 3.01 (0.962–9.42) | 1.67 (0.497–5.64) | 2.05 (0.877–4.8) |
| Education | | | |
| *Primary* | 1.94 (0.95–3.96) | 1.52 (0.828–2.78) | 1.61 (0.999–2.59) |
| *Secondary* | 1.29 (0.694–2.41) | 1.52 (0.742–3.12) | 1.44 (0.89–2.34) |
| *Tertiary* | 3.45 (1.748–6.81) | 2.15 (0.944–4.91) | 2.64 (1.531–4.55) |
| BMI | | | |
| *Underweight* | - - | - - | - - |
| *Normal* | 1.3 (0.705–2.38) | 1.02 (0.655–1.59) | 1.09 (0.707–1.69) |
| *Overweight* | 2.18 (1.38–3.45) | 1.65 (1.148–2.37) | 1.83 (1.348–2.47) |
| *Obese* | 1.13 (0.533–2.38) | 1.06 (0.558–1.99) | 1.07 (0.58–1.98) |

CU, cognitively unimpaired; MCI, mild cognitive impairment; RR, risk ratio; CI, confidence interval; *APOE*, apolipoprotein E; BMI, body mass index. RRs are given using the low neurofilament light (NfL) range as reference. Estimates were obtained from a quasi-Poisson regression model with a log link, adjusted for age, sex, education, *APOE* genotype, and BMI, incorporating interaction terms between NfL range, cognitive status and characteristics at HUNT4 70+. No significant differences between sexes were found when evaluated by rate of risk ratios tests. Inverse probability weighting was applied to reduce bias due to selective participation. Plasma NfL data were missing from 63 participants. Two participants had cognitive impairment other than MCI.

#### Supplementary tables on joint p-tau217 and NfL and AD dementia

Table S34. Estimated absolute risk of clinical Alzheimer's disease dementia by baseline cognitive status, joint plasma p-tau217 and NfL range and sex.

| Clinical Diagnosis at baseline | Plasma p-tau217 and NfL at baseline | Female | | | Male | | | Total | | |
|---|---|---|---|---|---|---|---|---|---|---|
| | | N | Risk (95% CI) | | N | Risk (95% CI) | | N | Risk (95% CI) | |
| CU | < cut-off | 935 | 3.6 | (2.5–5.3) | 680 | 2.9 | (1.8–4.7) | 1615 | 3.3 | (2.5–4.5) |
| | ≥ cut-off | 55 | 14.9 | (7.3–28.2) | 61 | 23.5 | (13.5–37.6) | 116 | 19.2 | (12.5–28.4) |
| | All | 1746 | 6.6 | (5.4–8.0) | 1424 | 5.5 | (4.3–7.0) | 3170 | 6.1 | (5.2–7.1) |
| MCI | < cut-off | 398 | 16.3 | (12.5–21.0) | 392 | 11.9 | (8.8–16.0) | 790 | 14.2 | (11.6–17.2) |
| | ≥ cut-off | 52 | 48.9 | (34.6–63.5) | 49 | 42.2 | (27.8–58.0) | 101 | 46 | (35.7–56.7) |
| | All | 894 | 25.4 | (22.2–28.8) | 905 | 18.7 | (16.1–21.8) | 1799 | 22.2 | (20.1–24.5) |
| Total | < cut-off | 1333 | 7.9 | (6.4–9.9) | 1072 | 6.6 | (5.1–8.5) | 2405 | 7.4 | (6.2–8.7) |
| | ≥ cut-off | 107 | 33.8 | (24.6–44.4) | 110 | 32.8 | (23.7–43.4) | 217 | 33.3 | (26.7–40.7) |
| | All | 2642 | 14.0 | (12.5–15.7) | 1147 | 11.2 | (9.8–12.8) | 3789 | 12.7 | (11.7–13.9) |

p-tau217, phosphorylated tau at threonine 217; NfL, neurofilament light; CI, confidence interval; CU, cognitively unimpaired; MCI, mild cognitive impairment; Plasma p-tau217 data was missing from one participant. Plasma NfL data were missing from 35 females and 28 males. Two participants had cognitive impairment other than MCI. Estimates are weighted.

Table S35. Estimated absolute risk of clinical Alzheimer's disease dementia by baseline cognitive status, joint plasma p-tau217 and NfL ranges and age groups.

| Clinical Diagnosis at baseline | Plasma p-tau217 and NfL at baseline | Age, 70–74 | | | Age, 75–79 | | | Age, 80–84 | | | Age, 85–89 | | | Age, 90+ | | |
|---|---|---|---|---|---|---|---|---|---|---|---|---|---|---|---|---|
| | | N | Risk (95% CI) | | N | Risk (95% CI) | | N | Risk (95% CI) | | N | Risk (95% CI) | | N | Risk (95% CI) | |
| CU | < cut-off | 966 | 2.3 | (1.4–3.6) | 457 | 3.3 | (1.9–5.8) | 154 | 5.8 | (3.0–10.9) | 29 | 10.4 | (3.2–29.3) | 9 | 17.9 | (3.3–58.5) |
| | ≥ cut-off | 27 | 10.8 | (2.5–37.0) | 24 | 11.9 | (3.6–32.9) | 32 | 21.0 | (9.7–39.8) | 23 | 27.0 | (11.7–50.9) | 10 | 21.7 | (4.1–64.2) |
| | All | 1635 | 3.4 | (2.5–4.5) | 956 | 5.4 | (4.0–7.1) | 426 | 10.5 | (7.8–14.0) | 124 | 16.8 | (10.9–25.0) | 29 | 18.8 | (8.1–37.9) |
| MCI | < cut-off | 476 | 8.1 | (5.8–11.2) | 235 | 19.8 | (14.8–26.0) | 67 | 18.2 | (10.2–30.3) | 8 | 55.4 | (18.0–87.6) | 4 | 72.3 | (5.8–99.1) |
| | ≥ cut-off | 21 | 65.6 | (42.2–83.3) | 24 | 18.0 | (6.5–41.1) | 29 | 49.2 | (30.3–68.4) | 18 | 38.4 | (17.3–65.0) | 9 | 79.9 | (38.2–96.3) |
| | All | 963 | 12.6 | (10.5–15.1) | 531 | 22.0 | (18.4–26.0) | 226 | 33.0 | (26.7–40.0) | 84 | 45.6 | (34.7–57.0) | 22 | 61.2 | (37.7–80.5) |
| Total | < cut-off | 1442 | 4.4 | (3.4–5.8) | 692 | 9.6 | (7.3–12.4) | 221 | 10.1 | (6.6–15.3) | 37 | 22.8 | (10.8–41.8) | 13 | 40.0 | (14.9–71.8) |
| | ≥ cut-off | 48 | 37.8 | (24.0–53.8) | 48 | 15.3 | (7.2–29.7) | 61 | 35.6 | (23.6–49.7) | 41 | 33.1 | (19.2–50.7) | 19 | 54.0 | (30.1–76.2) |
| | All | 2572 | 7.1 | (6.1–8.3) | 1488 | 12.0 | (10.3–14.0) | 652 | 19.6 | (16.4–23.3) | 208 | 31.2 | (24.7–38.5) | 51 | 41.0 | (27.4–56.2) |

p-tau217, phosphorylated tau at threonine 217; NfL, neurofilament light; CI, confidence interval; CU, cognitively unimpaired; MCI, mild cognitive impairment; Age is given in years. Plasma p-tau217 data was missing from one participant. Plasma NfL data were missing from 63 participants. Two participants had cognitive impairment other than MCI. Estimates are weighted.

Table S36. Estimated absolute risk of clinical Alzheimer's disease dementia by baseline cognitive status, joint plasma p-tau217 and NfL range and *APOE* genotype.

| **Clinical Diagnosis at baseline** | **Plasma p-tau217 and NfL at baseline** | **ε3ε3/ε3ε2/ε2ε2** | | | **ε2ε4/ε3ε4** | | | **ε4ε4** | | |
|---|---|---|---|---|---|---|---|---|---|---|
| | | **N** | **Risk** | **(95% CI)** | **N** | **Risk** | **(95% CI)** | **N** | **Risk** | **(95% CI)** |
| CU | < cut-off | 1309 | 3.5 | (2.5–4.8) | 286 | 3.1 | (1.5–6.2) | 14 | - | - |
| | ≥ cut-off | 72 | 9.4 | (4.4–19.0) | 40 | 33.7 | (19.3–51.9) | 4 | 58.6 | (5.4–97.2) |
| | All | 2293 | 5.5 | (4.5–6.7) | 811 | 7.0 | (5.2–9.2) | 55 | 16.5 | (8.6–29.5) |
| MCI | < cut-off | 641 | 14.9 | (12.0–18.3) | 142 | 10.8 | (6.1–18.3) | 2 | - | - |
| | ≥ cut-off | 61 | 45.3 | (32.2–59.0) | 35 | 47.1 | (29.8–65.1) | 5 | 49.5 | (6.4–93.4) |
| | All | 1226 | 20.8 | (18.3 -23.5) | 503 | 23.9 | (19.9–28.4) | 54 | 36.9 | (23.9–52.1) |
| Total | < cut-off | 1950 | 7.6 | (6.4–9.1) | 428 | 6.0 | (3.8–9.3) | 16 | - | - |
| | ≥ cut-off | 133 | 28.7 | (20.7–38.2) | 75 | 40.4 | (28.9- 53.1) | 9 | 53.3 | (18.7 -85.0) |
| | All | 3520 | 11.5 | (10.3–12.9) | 1315 | 14.4 | (12.3–16.8) | 109 | 27.3 | (19.0–37.6) |

p-tau217, phosphorylated tau at threonine 217; NfL, neurofilament light; *APOE*, apolipoprotein E; CI, confidence interval; CU, cognitively unimpaired; MCI, mild cognitive impairment; 2 participants had cognitive impairment other than MCI. Plasma p-tau217 data was missing from one participant. Plasma NfL data were missing from 63 participants. *APOE* genotype data was missing from 11 CU and 16 MCI participants. Estimates are weighted.

Table S37. Estimated absolute risk of clinical Alzheimer's disease dementia by baseline cognitive status, plasma p-tau217 and NfL range and educational level.

| Clinical Diagnosis at baseline | Plasma p-tau217 and NfL at baseline | Primary education | | | Secondary education | | | Tertiary education | | |
|---|---|---|---|---|---|---|---|---|---|---|
| | | N | Risk (95% CI) | | N | Risk (95% CI) | | N | Risk (95% CI) | |
| CU | < cut-off | 307 | 4.8 | (2.9–8.0) | 732 | 3.7 | (2.5–5.4) | 576 | 1.3 | (0.6–2.8) |
| | ≥ cut-off | 31 | 17.7 | (7.3–36.9) | 42 | 23.3 | (12.3–39.8) | 43 | 16 | (7.2–31.7) |
| | All | 646 | 7.7 | (5.8–10.2) | 1426 | 6.3 | (5.1–7.8) | 478 | 3.8 | (2.8–5.2) |
| MCI | < cut-off | 204 | 20.7 | (15.4–27.4) | 382 | 12.5 | (9.5–16.4) | 204 | 5 | (2.6–9.4) |
| | ≥ cut-off | 29 | 60.1 | (40.5–77.0) | 49 | 34.5 | (22.0–49.6) | 22 | 44.2 | (24.3–66.1) |
| | All | 481 | 30.2 | (26.0–34.8) | 863 | 18.7 | (16.1–21.7) | 452 | 12.5 | (9.6–16.1) |
| Total | < cut-off | 511 | 11.7 | (9.0–15.2) | 1114 | 7.0 | (5.5–8.7) | 780 | 2.4 | (1.4–3.8) |
| | ≥ cut-off | 60 | 40.7 | (28.3–54.3) | 91 | 29.9 | (20.9–40.7) | 65 | 26.4 | (16.7–39.1) |
| | All | 1127 | 18.5 | (16.1–21.1) | 2290 | 11.4 | (10.1–12.9) | 1550 | 6.6 | (5.4–8.0) |

p-tau217, phosphorylated tau at threonine 217; NfL, neurofilament light; CI, confidence interval; CU, cognitively unimpaired; MCI, mild cognitive impairment; Two participants had cognitive impairment other than MCI. Plasma p-tau217 data was missing from one participant. Plasma NfL data were missing from 63 participants. Education status was missing from four participants. Estimates are weighted. "Primary education" was defined ≤10 years of compulsory primary and lower secondary education; "Secondary education" was defined as additional 1–2 years of academic or vocational school, 3 years of academic or vocational school, or 3–4 years of vocational training or apprenticeship (upper secondary education); "Tertiary education" referred to college or university education.

Table S38. Estimated absolute risk of clinical Alzheimer's disease dementia by baseline cognitive status, plasma p-tau217 and NfL range and body mass index.

| Clinical Diagnosis at baseline | Plasma p-tau217 and NfL at baseline | BMI, Underweight | | | BMI, Normal | | | BMI, Overweight | | | BMI, Obese | | |
|---|---|---|---|---|---|---|---|---|---|---|---|---|---|
| | | N | Risk (95% CI) | | N | Risk (95% CI) | | N | Risk (95% CI) | | N | Risk (95% CI) | |
| CU | < cut-off | 6 | - | - | 446 | 2.0 | (0.9–4.4) | 771 | 3.9 | (2.6–5.6) | 377 | 2.5 | (1.3–4.9) |
| | ≥ cut-off | 0 | - | - | 41 | 11.8 | (4.7–26.9) | 44 | 28.5 | (16.1–45.4) | 25 | 13.1 | (3.7–36.7) |
| | All | 14 | 12.4 | (2.6–42.9) | 937 | 5.6 | (4.2–7.5) | 1512 | 6.4 | (5.2–8) | 678 | 5.1 | (3.5–7.3) |
| MCI | < cut-off | 2 | - | - | 181 | 12.7 | (8–19.5) | 380 | 12 | (8.7–16.5) | 222 | 17.9 | (13–24.1) |
| | ≥ cut-off | 0 | - | - | 29 | 48.0 | (29–67.6) | 45 | 44.4 | (29.4–60.4) | 14 | 18.6 | (4.9–50.4) |
| | All | 13 | 21.6 | (5.7–55.6) | 491 | 24.3 | (20.3–29) | 801 | 19.0 | (16–22.4) | 439 | 20.8 | (16.9–25.4) |
| Total | < cut-off | 8 | - | - | 627 | 5.5 | (3.7–8.1) | 1151 | 6.9 | (5.4–8.9) | 599 | 8.7 | (6.5–11.6) |
| | ≥ cut-off | 0 | - | - | 70 | 27.8 | (17.5–41.1) | 89 | 37.6 | (27.3–49.2) | 39 | 15.2 | (6.5–31.8) |
| | All | 27 | 17.1 | (6.6–37.5) | 1428 | 13.0 | (11–15.2) | 2315 | 11.4 | (10–13.1) | 1117 | 11.8 | (9.8–14.2) |

p-tau217, phosphorylated tau at threonine 217; NfL, neurofilament light; BMI, body mass index; CI, confidence interval; CU, cognitively unimpaired; MCI, mild cognitive impairment. Two participants had cognitive impairment other than MCI. Plasma p-tau217 data was missing from one participant. Plasma NfL data were missing from 63 participants. BMI data were missing from 29 CU participants and 55 MCI participants. Estimates are weighted.

Table S39. Association between baseline concordant high versus low p-tau217 and NfL ranges and risk of clinical Alzheimer's disease dementia at follow-up across demographic and clinical subgroups (N, 2,572).

| **Characteristics at baseline** | **CU** | **MCI** | **Total** |
|---|---|---|---|
| | **RR (95% CI)** | **RR (95% CI)** | **RR (95% CI)** |
| All | 3.28 (1.53–7.03) | 3.75 (2.54–5.53) | 3.63 (2.57–5.14) |
| Sex | | | |
| *Female* | 3.04 (1.2–7.72) | 3.04 (1.94–4.78) | 3.04 (2.01–4.59) |
| *Male* | 3.62 (1.41–9.27) | 4.71 (2.68–8.28) | 4.44 (2.73–7.22) |
| Age | | | |
| *70–74* | 4.29 (1.11–16.58) | 10.59 (6.295–17.81 | 8.58 (5.33–13.83) |
| *75–79* | 2.61 (0.776–8.75) | 1.18 (0.432–3.24) | 1.47 (0.681–3.19) |
| *80–84* | 2.77 (0.945–8.12) | 2.51 (1.221–5.15) | 2.6 (1.417–4.78) |
| *85–89* | 3.33 (0.706–15.66) | 0.64 (0.253–1.64) | 1.07 (0.514–2.24) |
| *90+* | 1.36 (0.114–16.17) | 1.48 (0.615–3.58) | 1.47 (0.636–3.38) |
| *APOE* ε4 allele | | | |
| *ε3ε3/ε3ε2/ε2ε2* | 2.32 (0.923–5.8) | 3.57 (2.26–5.64) | 3.26 (2.17–4.91) |
| *ε2ε4/ε3ε4* | 6.30 (2.17–18.3) | 4.52 (2.48–8.25) | 5.05 (2.92–8.75) |
| *ε4ε4* | - - | - - | - - |
| Education | | | |
| *Primary* | 2.84 (1.076–7.5) | 3.58 (2.161–5.93) | 3.44 (2.19–5.4) |
| *Secondary* | 2.64 (0.909–7.66) | 2.82 (1.542–5.15) | 2.76 (1.63–4.68) |
| *Tertiary* | 7.35 (2.377–22.73) | 11.05 (4.898–24.94 | 9.61 (5–18.48) |
| BMI | | | |
| *Underweight* | - - | - - | - - |
| *Normal* | 3.32 (1.553–7.09) | 3.47 (2.379–5.07) | 3.44 (2.45–4.83) |
| *Overweight* | 3.38 (1.56–7.31) | 3.36 (2.242–5.05) | 3.37 (2.336–4.86) |
| *Obese* | 2.96 (1.349–6.51) | 4.64 (3.098–6.96) | 4.3 (2.985–6.2) |

p-tau217, phosphorylated tau at threonine 217; NfL, neurofilament light; CU, cognitively unimpaired; MCI, mild cognitive impairment; RR, risk ratio; CI, confidence interval; *APOE*, apolipoprotein E; BMI, body mass index. RRs are given using the low p-tau217 and NfL ranges as reference. Estimates were obtained from a quasi-Poisson regression model with a log link, adjusted for age, sex, education, *APOE* genotype, and BMI, incorporating interaction terms between p-tau217 and NfL ranges, cognitive status and characteristics at HUNT4 70+. No significant differences between sexes were found when evaluated by rate of risk ratios tests. Inverse probability weighting was applied to reduce bias due to selective participation. Plasma p-tau217 data was missing from one participant. Plasma NfL data were missing from 63 participants. Two participants had cognitive impairment other than MCI.

#### Data availability statement

To protect participants' privacy, HUNT Research Centre aims to limit storage of data outside HUNT databank and cannot deposit data in open repositories. HUNT databank has precise information on all data exported to different projects and can reproduce them on request. There are no restrictions regarding data export given approval of applications to HUNT Research Centre. Researchers can apply for data at https://www.ntnu.edu/hunt/research
